# Sociodemographic, clinical, and genetic factors associated with self-reported antidepressant response outcomes in the UK Biobank

**DOI:** 10.1101/2024.10.23.24315970

**Authors:** Michelle Kamp, Chris Wai Hang Lo, Grigorios Kokkinidis, Mimansa Chauhan, Alexandra C. Gillett, AMBER Research Team, Andrew M. McIntosh, Oliver Pain, Cathryn M. Lewis

## Abstract

**Background:** In major depressive disorder (MDD), only ∼35% achieve remission after first-line antidepressant therapy. Using UK Biobank data, we identify sociodemographic, clinical, and genetic predictors of antidepressant response through self-reported outcomes, aiming to inform personalized treatment strategies.

**Methods:** In UK Biobank Mental Health Questionnaire 2, participants with MDD reported whether specific antidepressants helped them. We tested whether retrospective lifetime response to four selective serotonin reuptake inhibitors (SSRIs) (N=19,516)-citalopram (N=8,335), fluoxetine (N=8,476), paroxetine (N=2,297) and sertraline (N=5,883)-was associated with sociodemographic (e.g., age, gender) and clinical factors (e.g., episode duration). Genetic analyses evaluated the association between CYP2C19 variation and self-reported response, while polygenic score (PGS) analysis assessed whether genetic predisposition to psychiatric disorders and antidepressant response predicted self-reported SSRI outcomes.

**Results:** 71%-77% of participants reported positive responses to an SSRI. Non-response was significantly associated with self-medication (OR=1.56, p=2.08×10^−19^), younger age (OR=0.87, p=2.49×10^−11^), male gender (OR=1.26, p=3.18×10^−08^), and lower income (OR=1.34, p=6.17×10^−07^). A worst episode lasting over two years (OR=1.97, p=1.25×10^−16^) and no mood improvement from positive events (OR=1.33, p=1.35×10^−06^) were also associated with non-response. CYP2C19 poor metabolizers had higher non-response rates (OR=1.31, p=1.77×10^−02^). Additionally, higher PGS for depression (OR=1.08, p=3.37×10^−05^), ADHD (OR=1.06, p=1.06×10^−03^), and schizophrenia (OR =1.06, p=1.53×10^−03^) predicted negative SSRI outcomes.

**Conclusion:** Self-reported antidepressant response in the UK Biobank is influenced by sociodemographic, clinical, and genetic factors, mirroring clinical response measures. While positive outcomes are more frequent than remission reported in clinical trials, these self-reports replicate known treatment associations, suggesting they capture meaningful aspects of antidepressant effectiveness from the patient’s perspective.

## INTRODUCTION

Major depressive disorder (MDD) is a prevalent and debilitating condition characterized by persistent low mood, loss of interest, cognitive impairment, and physical symptoms such as disrupted sleep or appetite (Otte et al., 2016). Affecting approximately one in six adults globally, MDD incidence continues to rise annually (Abdoli et al., 2022; GBD 2019 Mental Disorders Collaborators, 2022). In 2018, the World Health Organization (WHO) ranked MDD third in global disease burden and predicted it will be the leading cause by 2030 (Cui et al., 2024).

Antidepressants, specifically selective serotonin reuptake inhibitors (SSRIs), are first-line pharmacological treatments for MDD (Bauer, Severus, Möller, & Young, 2017; Cleare et al., 2015; NICE, 2009). In England’s National Health Service (NHS), antidepressant prescriptions nearly doubled from 36 million in 2008 to 70.9 million in 2018 (Iacobucci, 2019). SSRIs have similar or greater efficacy than other antidepressants and are preferred clinically for their fewer side effects (Cipriani et al., 2018; Karrouri, Hammani, Benjelloun, & Otheman, 2021). However, antidepressant efficacy varies with only ∼35% of patients achieving remission after initial treatment (Rush et al., 2006), and approximately 40% developing treatment-resistant depression, defined as the lack of response to two or more antidepressants in the same depressive episode (Rush et al., 2011; Souery et al., 2011).

Variability in treatment response may be due to disorder heterogeneity (Cui et al., 2024; Fried & Nesse, 2015), genetics (Pain et al., 2022; Tansey et al., 2013), and sociodemographic or clinical factors (Perna, Alciati, Daccò, Grassi, & Caldirola, 2020). While studies assessing the impact of age and sex on antidepressant response show inconsistent results (Kessler et al., 2017; Khan, Brodhead, Schwartz, Kolts, & Brown, 2005; Perna et al., 2020; Saveanu et al., 2015; Trivedi et al., 2006); socioeconomic factors such as low income and unemployment have been associated with poor response to the antidepressant citalopram (Trivedi et al., 2006). Genome-wide association studies (GWASs) have yet to find robust genetic predictors of antidepressant response, likely due to small sample sizes, study design and drug and outcome heterogeneity (Ising et al., 2009; Ji et al., 2013; Uher et al., 2009). Nevertheless, SNP-based heritability estimates by the Psychiatric Genomics Consortium suggest 13% to 40% of the variance in antidepressant response is attributable to common genetic variation (Pain et al., 2022).

Pharmacogenetic studies have identified variation in the cytochrome P450 (CYP) enzyme superfamily that affects drug response via pharmacokinetic mechanisms. Polymorphisms in *CYP* genes, including *CYP2C19*, impact enzyme activity and may explain individual differences in treatment response (Li et al., 2024; Wong et al., 2023). However, CYP2C19 and other candidate genes account for only a small fraction of variability in drug response. Polygenic scores (PGS) offer an alternative by quantifying an individual’s genetic predisposition to a trait or disease, aggregating effects of multiple SNPs identified through GWAS. By capturing the polygenic nature of treatment response, where many loci contribute small effects, PGS may be valuable for predicting response. Although PGS for bipolar disorder and MDD, based on relatively small sample sizes, have shown inconsistent associations with treatment outcomes (Fanelli et al., 2021; García-González et al., 2017), positive antidepressant response has been associated with low genetic liability for schizophrenia (Pain et al., 2022).

The trial-and-error approach to finding the right antidepressant delays recovery, increases side effects, reduces adherence, and highlights the need to identify individual moderators of treatment response to support personalized treatments (Perna et al., 2020). Most studies identifying factors associated with antidepressant response come from clinical trials, which often have limited generalizability due to restrictive inclusion criteria and controlled settings. However, comprehensive datasets, including retrospective self-report on antidepressant response, are becoming available (Koch et al., 2024). Little is known about predictors associated with an SSRI-user reporting that an antidepressant ‘helped’ them. This study uses retrospective self-report data from approximately 20,000 UK Biobank participants to assess sociodemographic, clinical, and genetic predictors of this patient-focused measure of SSRI response and compare them with the those identified in prospective clinical studies.

## METHOD

### Participants

The UK Biobank (UKB) is a large-scale research resource investigating the impact of genetic, environmental, and lifestyle factors on health outcomes in middle-aged and older adults. Over 9,000,000 individuals aged 40 to 69 registered with the UK National Health Service were invited to participate. Of those, ∼500,000 individuals were recruited in 22 assessment centers across the UK between 2006 and 2010 (Allen, Sudlow, Peakman, & Collins, 2014). Baseline data included sociodemographic characteristics, medical histories, and health and lifestyle factors. During follow-up (2016–2017), 157,270 participants completed the online mental health questionnaire (MHQ). A subsequent follow-up questionnaire (MHQ2), focusing on mental health and wellbeing (Category 1502), was initiated in 2022 (N=172,912). Participants who endorsed at least one of two cardinal lifetime MDD symptom screening questions (UKB field 29011 and 29012, Supplementary materials1, Table S1) completed the medication and antidepressant response sections (N=79,888). After excluding participants with prior or probable schizophrenia, bipolar, or mania diagnoses (N=1,527, from UKB fields 29000 and 20126), 78,361 eligible participants remained.

Prescription medication users (Yes/No) (N=35,956) were identified from those who reported trying prescribed medication for low mood or anhedonia (UKB field 29038, Supplementary materials1, TableS1). Antidepressant users were prescription medication users who reported trying specific antidepressants for at least two weeks (UKB field 29039), including citalopram, fluoxetine, sertraline, paroxetine, amitriptyline, dosulepin, or other antidepressant(s) (N=28,673). SSRI users were those who had tried at least one SSRI (citalopram, fluoxetine, paroxetine, or sertraline) (N=21,124) (Supplementary Materials1, FigureS1). Association analyses were restricted to SSRI users who answered “Yes” or “No” to whether the SSRI helped them feel better (UKB field 29040, 29041, 29042 and 29043), i.e. “Do not know” and “Prefer not to answer” were excluded (final SSRI N=19,516).

### Ethics and consent

The UK Biobank has research ethics approval from the North West Multi-center Research Ethics Committee (MREC; approval number 11/NW/0382) covering the UK. Participation is voluntary, and participants can withdraw at any time. Informed written consent was obtained at baseline. The current study was performed under UK Biobank application 82087. All relevant ethical guidelines have been followed during analysis.

### Measures

#### Self-reported antidepressant response outcomes

Antidepressant response was restricted to participants reporting “Yes (Y)” or “No (N)” to trying at least one of the following SSRIs for at least two weeks: citalopram, fluoxetine, paroxetine, sertraline (N=19,516).

##### Drug-specific SSRI antidepressant response

Drug-specific SSRI response outcomes were binary, with a positive response defined as “Y” to where users had responded “Yes, even a little,” to whether the specific SSRI (citalopram, fluoxetine, paroxetine, or sertraline) they tried helped them “feel better”. The drug-specific response was viewed in isolation, regardless of other self-reported antidepressant exposures. The reference category for drug-specific SSRI response outcomes was positive response (Y).

##### Overall SSRI antidepressant response

In addition to the four drug-specific outcomes, a binary composite-SSRI response outcome was also considered. For participants with responses to multiple SSRIs, a decision framework based on self-reported outcomes for each antidepressant was used to define their composite-SSRI response phenotype. Participants with consistent responses across antidepressants (i.e. all “Y”, or all “N”) were categorized “Y” or “N” accordingly. For any participant reporting “N” for any antidepressant their composite-SSRI response was “N”. A binary SSRI-conservative outcome was also defined for those using multiple SSRIs, assigning a missing value where only one “N” was reported (Supplementary materials1, TableS2). The reference category for both overall SSRI responses was a positive response.

#### Sociodemographic factors

Sociodemographic variables collected at baseline UKB assessment were analyzed, including age, sex, ethnicity, educational attainment, household income, and neighborhood deprivation. Age at recruitment was recorded in years. Gender was self-reported as biological sex (female/male). Ethnic background was self-reported as “Asian or Asian British”, “Black or Black British”, “Mixed background”, “White” and “Other ethnic group”; for analysis, categories were grouped as “Asian,” “Black,” “Mixed,” and “White,” with White as the reference.

Educational attainment was classified into six categories based on highest education level achieved (Rayner et al., 2021): ‘Secondary’ for completion of compulsory secondary education (GCSE level, 11 years of education); ‘Further’ for completion of further education (A-levels, 13years); ‘Vocational’ for a range of vocational and professional qualifications (NVQs, BTECs, Apprenticeships, ≥12years); and ‘University’ for university-level education (Degree, ≥16years). As the modal category, University was set as the reference. Annual household income was divided into five categories: Less than £18,000, £18,000-£31,000, £31,000-£52,000, £52,000-£100,000, and Greater than £100,000, with the median category (£30,000-£52,000) serving as the reference.

Neighborhood deprivation was measured using the Townsend Deprivation Index (TDI), which combines four census variables (unemployment, car ownership, homeownership, household overcrowding), standardized and summed to a total score. Areas with TDI>0 are more deprived, while TDI<0 indicates more affluent areas. A variable for self-medication with drugs and alcohol was created based on participants’ reports of using alcohol or drugs to manage their two cardinal MDD symptoms as these coping mechanisms have linked to treatment seeking behavior for MDD (Rayner et al., 2021).

#### Clinical factors

MDD symptoms during a participant’s worst episode of MDD are available for UKB participants who answered relevant questions derived from the Composite International Diagnostic Interview (CIDI) conducted through the MHQ2, data category 1502 (World Health Organization, 1993). The questions identify key depression symptoms, including persistent sadness, loss of interest or pleasure, changes in weight or sleep, fatigue, guilt, worthlessness, concentration difficulties, and suicidal ideation (World Health Organization, 1993).

The MHQ2 also assessed clinical characteristics of MDD through self-report, including the lifetime number of depressive periods, age at first and last episode, and whether episodes were related to childbirth or trauma. Family history of severe depression was determined if the participant reported their father, mother or sibling experienced severe depression (UKB field 20107, 20110, 20111); as the modal category, “No” was the reference category. The full list of clinical variables, their data fields, and references categories used in this analysis are in Supplementary materials1, TableS3.

#### Genetic factors

##### CYP2C19 metabolizer status

CYP2C19 is a key enzyme in SSRI metabolism, with genetic variation in the CYP2C19 gene associated with differential metabolic activity and, consequently, differential SSRI exposure. Metabolic capacity is determined by specific allelic variants, with patient classified into one of five metabolizer groups based on genotypes and corresponding enzymatic function - poor (no functional enzyme), intermediate (reduced enzyme activity), normal (normal enzyme activity), rapid, and ultra-rapid (increased enzyme activity). CYP2C19 genotypes and metabolizer status were obtained from UKB return 3388 as described by McInnes et al. (McInnes et al., 2021). In brief, pharmacogenetic star alleles and metabolizer phenotypes were identified using the Python program PGxPOP, which determines and reports pharmacogenetic star alleles (popular nomenclature that corresponds to functional haplotype patterns which influence drug metabolism) and haplotypes from phased multisample VCFs using PharmCAT allele definition files (https://github.com/PharmGKB/PharmCAT) (Sangkuhl et al., 2020). PGxPOP applies guidelines from the Dutch Pharmacogenetics Working Group (DPWG) and the Clinical Pharmacogenetics Implementation Consortium (CPIC) to correlate haplotypes with predicted metabolic phenotypes. Participants were categorized into five metabolic phenotypes—poor (PM), intermediate (IM), normal (NM), rapid (RM) and ultrarapid (UM)—based on predicted CYP2C19 activity. Individuals with “indeterminate” or “not available” phenotypes were excluded due to unknown, uncertain, or unaligned star allele functions, preventing phenotype assignment.

##### Polygenic scores

Polygenic scores (PGS) were calculated for five psychiatric disorders and two antidepressant response outcomes using GWAS summary statistics from the Psychiatric Genomics Consortium (Demontis et al., 2019; Grove et al., 2019; Pain et al., 2022; Pardiñas et al., 2018; Stahl et al., 2019; Wray et al., 2018) (Supplementary materials1, TableS4). The psychiatric disorders included depression (DEPR), autism (AUTI), Attention-Deficit/Hyperactivity Disorder (ADHD), bipolar disorder (BIPO), and schizophrenia (SCHI). The two antidepressant response outcomes were percentage improvement (ADperc), and non-remission (ADnon-rem) (Pain et al., 2022). ADperc was calculated as 100*(baseline depression score – final depression score)/baseline depression score, with higher ADperc indicating better treatment response. Remission is a binary measure attained when a patient’s depression symptom score decreased to a pre-specified threshold for the relevant rating scale. Patients who did not reach these thresholds were classified as non-remitting (Pain et al., 2022).

PGS were calculated using the MegaPRS method within the GenoPred Pipeline (https://opain.github.io/GenoPred/) (Pain, Al-Chalabi, & Lewis, 2024). The MegaPRS model selected by its pseudo summary approach was used in downstream analyses. The pseudo summary approach estimates the best tuning parameters based solely on the GWAS summary statistics, avoiding the need for an external validation sample. MegaPRS was used due to its superior prediction performance for psychiatric disorders (Ni et al., 2021). Covariates included the first six principal components (to adjust for population stratification), age, sex, and genotyping batch. Assessment Center was excluded as a covariate as association testing showed no significant association with self-reported response outcomes (Supplementary Materials1, TableS5).

### Statistical analyses

Six outcome measures were analyzed: self-reported binary response (Y/N) for four SSRIs (citalopram, fluoxetine, paroxetine, or sertraline), overall-SSRI response, and conservative-SSRI response. Separate association analyses were conducted to assess associations between sociodemographic, clinical, and genetic predictors with antidepressant response (two SSRI responses and drug-specific) using univariable and multivariable logistic regression models and the odds ratio (OR) reported. The reference level was a positive response; hence, all ORs describe the odds of SSRI non-response compared to positive response. To address potential sex differences in antidepressant response, gender-stratified analyses were conducted. In gender-stratified sociodemographic analyses ethnic background was removed due to small sample sizes of minority groups. Given the large number of clinical predictors, only those reaching nominal significance in univariable analyses were taken forward into multivariable models. Variance inflation factors assessed multicollinearity in multivariable models (Supplementary materials1, Table S9-S10) and covariates included age and sex. Clinical and genomic analyses were limited to White Western European ancestry participants due to insufficient sample sizes in other ancestries.

All analyses were conducted using R (version 4.2.2). Logistic regressions were performed using the *glm* function (R Core Team, 2021). Associations between PGS and self-reported antidepressant response were reported as the OR for the standardized polygenic score. A Bonferroni correction accounted for the number of predictors in each multivariable model. A stricter correction was also applied to adjust for both the number of predictors and antidepressant response phenotypes (four drug-specific, composite-SSRI, and SSRI-conservative). Thresholds were: sociodemographic (eight factors tested; α = 0.05/8 = 0.007; strict α = 0.05/48 = 0.001), clinical (15 factors tested; α = 0.05/15 = 0.003; strict α = 0.05/90 = 0.0006), PGS (seven PGS tested – five psychiatric disorders/traits and two antidepressant response phenotypes; α = 0.05/7 = 0.007; strict α = 0.05/42 = 0.001) and CYPC2C19 metabolizer status (single predictor tested; α = 0.05; strict α = 0.05/6 = 0.008). Figures display drug-specific and composite-SSRI phenotypes, as conservative-composite-SSRI analyses closely matched the composite-SSRI phenotype.

## RESULTS

### Characteristics

After excluding individuals missing genetic information, a total of 76,526 individuals were available for this analysis. 45.9% (N=35,088) of these used prescription medications to alleviate symptoms, and 36.6% (N=27,977) tried antidepressants. About one-quarter (26.9%, N=20,613) used at least one SSRI. Figure 1 presents the total number of SSRI and specific drug counts among SSRI users. Across groups, approximately 7% responded with “Do not know” and <1% with “Prefer not to answer” (Figure1); these individuals were excluded from further analysis. Final sample sizes were citalopram (N=8,335), fluoxetine (N=8,476), paroxetine (N=2,297), and sertraline (N=5,883) (Figure1). As participants could use multiple SSRIs, the count for SSRI in Figure1 exceeds the number of unique participants. To address this, a decision framework (Methods: 2.33) was applied, resulting in 19,516 unique individuals for the composite-SSRI phenotype and 18,170 individuals for the conservative-SSRI group. Over three-quarters (77.8%) of SSRI individuals tried a single SSRI for at least two weeks, with citalopram and fluoxetine being the most common (∼35% each), followed by sertraline (21%) and paroxetine (8%). Approximately 5% tried three or more of these SSRIs (Supplementary materials1, FigureS3).

**Figure 1:**
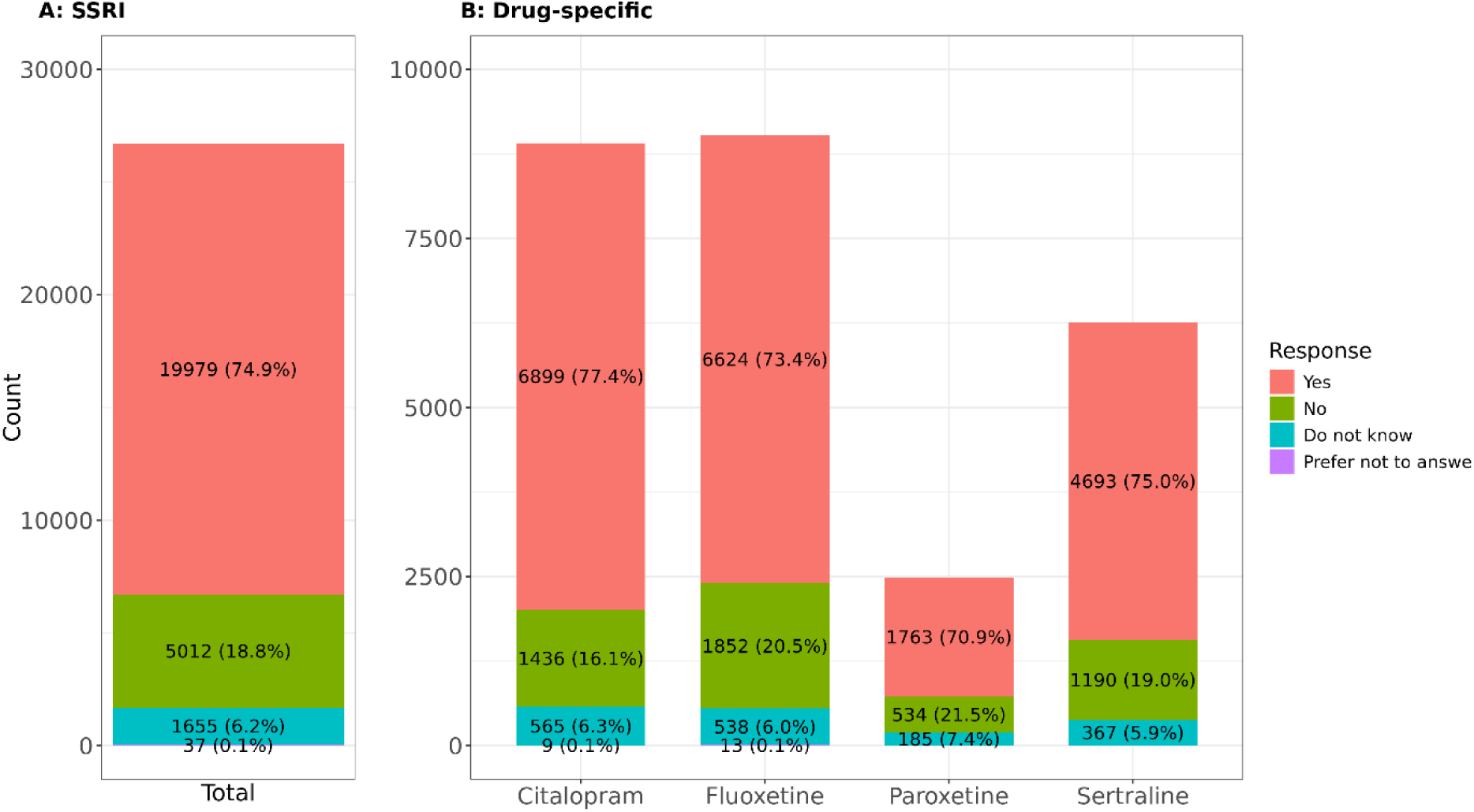
Distribution of total number of SSRI responses in a subset of UKB participants reporting at least one of the two cardinal symptoms for MDD. SSRI count does not match the number of participants taking SSRIs as some participants reported taking more than one antidepressant. Participants reported ‘Yes’ or ‘No’ that the SSRI drug helped them.

For participants who responded positively to at least one cardinal MDD question, we compared those who had (N=19,516) and had not (N=57,010) tried SSRIs. SSRI users were younger (55.0±7.5years vs 52.5±7.3years, *p*<1.00×10*^−300^*; Table2), more likely to be female (63.7 vs 73.6%, *p*=2.08×10^−142^) and of white ethnicity (97.9 vs 98.5%, *p*=6.18×10^−10^). SSRI users also had lower income (annual household income <£18 000: 13.9 vs 16.4%, *p*=1.50×10^−23^), came from less deprived neighborhoods (−1.6 vs −1.4, *p*=7.98×10^−11^) and were less likely to have a university degree (46.3 vs 43.8%, *p*=5.17×10^−18^).

**Table 2:** Participant characteristics among those who have and have not tried selective serotonin reuptake inhibitors (SSRI) in a subsample of the UKB participants reporting at least one of the two cardinal symptoms for MDD.

|  | SSRI not<br>tried | SSRI tried | *p value*<sup>1</sup> |
| --- | --- | --- | --- |
|  | N = 57 010 | N = 19 516 |  |
| Age |  |  |  |
| Mean (SD) | 55.0 (7.5) | 52.5 (7.3) | <1.00x10<sup>-300</sup> |
| Median | 56 | 52 |  |
| Range (min, max) | (40, 72) | (40, 70) |  |
| IQR | (49,61) | (47,58) |  |
| Sex |  |  |  |
| Female (%) | 36 287 (64) | 14 368 (74) | 2.08x10<sup>-142</sup> |
| Male (%) | 20 723 (36) | 5 148 (26) |  |
| Ethnic background |  |  |  |
| Asian (%) | 431 (0.8) | 81 (0.4) | 6.18x10<sup>-10</sup> |
| Black (%) | 375 (0.7) | 74 (0.4) |  |
| Mixed (%) | 361 (0.6) | 130 (0.7) |  |
| White (%) | 55 195 (98) | 19 081 (99) |  |
| Missing | 648 | 150 |  |
| Annual income |  |  |  |
| < £18 000 (%) | 7 145 (14) | 2 947 (16) | 1.50x10<sup>-23</sup> |
| £18 000 - £31 000 (%) | 12 012 (23) | 15 038 (23) |  |
| £31 000 - £52 000 (%) | 15 038 (29) | 5 399 (30) |  |
| £52 000 - £100 000 (%) | 13 509 (26) | 4 407 (25) |  |
| > £100 000 (%) | 3 780 (7) | 1 070 (6) |  |
| Missing | 5 526 | 1 510 |  |
| Educational attainment |  |  |  |
| Secondary (GCSEs, 11 years) | 7 896 (14) | 2 959 (15) | 5.17x10<sup>-18</sup> |
| Further (A-levels, 13 years) | 7 838 (14) | 2 940 (15) |  |
| Vocational (NVQs, BTECs, etc., ≥ 12 years) | 11 142 (20) | 3 995 (21) |  |
| University (Degree, ≥ 16 years) | 26 275 (46) | 8 506 (44) |  |
| None listed | 3 562 (6) | 1 020 (5) |  |
| Missing | 297 | 96 |  |
| Townsend Deprivation Index (TDI) |  |  |  |
| Mean (SD) | -1.6 (2.9) | -1.4 (3.0) | 7.98x10<sup>-11</sup> |
| Median | -2.3 | -2.2 |  |
| Range (min, max) | (-6.3, 10.5) | (-6.3, 11.0) |  |
| IQR | (-3.75, 0.05) | (-3.67, 0.35) |  |
| Missing | 74 | 29 |  |

<sup>1</sup> *p*-values were generated using the Pearson's chi-squared test for categorical variables and the Wilcoxon rank sum test for continuous variables; *p* < 0.05. GCSE - General Certificate of Secondary Education, A-levels - Advanced Level qualifications, NVQs - National Vocational Qualifications, BTECs - Business and Technology Education Council qualifications.

Drug-specific analysis showed consistent age and sex differences between those who had and had not tried specific SSRIs (e.g., Citalopram: Not tried vs. Tried, Supplementary materials2, TableS6). Participants who tried citalopram, fluoxetine, and sertraline were generally younger than those who had not tried the respective drug (citalopram: 52.9±7.3 vs 51.9±7.3years, *p*=5.1×10^−22^; fluoxetine: 52.8±7.3 vs. 52.1±7.1years, *p*=3.7×10^−12^; sertraline: 52.7±7.1 vs 51.9±7.5years, *p*=9.4×10^−16^); while paroxetine users were older (52.4±7.3 vs 53.2±7.2years, *p*=3.4×10^−08^). Females were more likely to try fluoxetine (72% vs 75%, *p*=3.5×10^−05^), while more males tried paroxetine (26% vs 30%, *p*=5.4×10^−03^) and sertraline (26% vs 28%, *p*=5.4×10^−03^).

Further demographic insights revealed fluoxetine users were more likely to be from less deprived neighborhoods and have a university degree (−1.5±3 vs −1.3±3, *p*=3.6×10^−03^; University degree: 43% vs 45%, *p*=5.6×10^−04^), while sertraline users were less educated (45% vs 41%, *p*=1.6×10^−04^), more likely to be non-white (White: 99% vs. 98%, *p*=0.02), and from lower-income groups (<£18K: 16% vs 18%). Paroxetine users had lower incomes (<£18K: 16% vs 18%, *p*=2.2×10^−04^) but were more educated (University degree: 43% vs 51%, *p*=3.2×10^−13^).

### Antidepressant exposure and response

Of the 20,613 individuals having tried at least one SSRI, 74.9% reported a positive response (“Y”), 18.8% indicated that the drugs did not work for them, ∼7% responded “Do not know” (∼7%) and <1% chose “Prefer not to answer” (<1%) (Figure2). When considering definitive responses only (“Y”/ “N”), 79.6% felt SSRIs made them “*feel better*” (Supplementary materials1, TableS7). This positive response was consistent across individual drugs, but rates varied by drug (χ²=73.28, *p*=8.5×10^−16^), highest for citalopram (82.8%) and lowest for paroxetine (76.8%). Sex-stratified analysis showed citalopram was more often linked to a positive outcome, and sertraline less so in females. In males, fluoxetine was less frequently associated with positive outcomes, while sertraline was more frequently associated (Supplementary materials1, TableS8).

**Figure 2:**
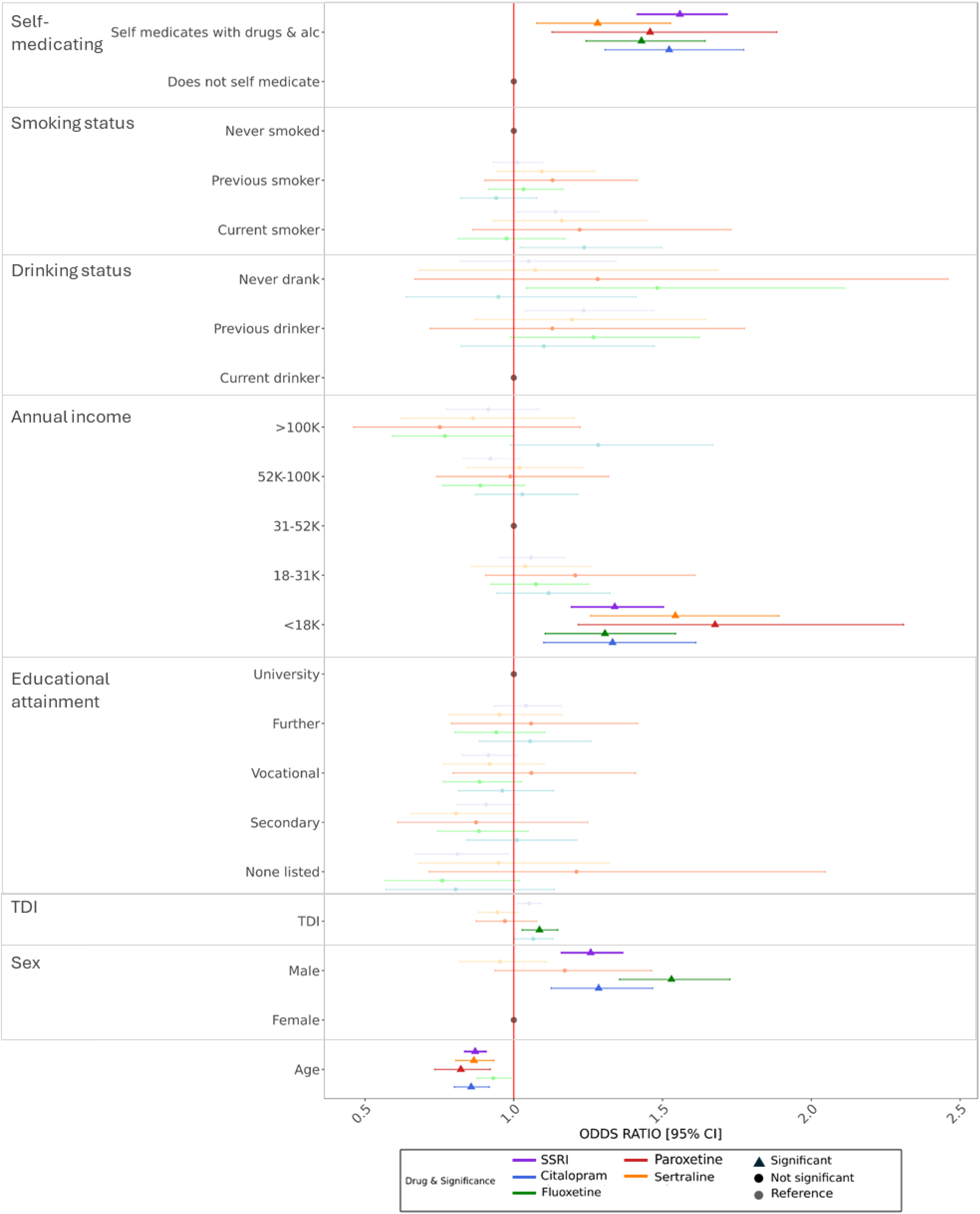
Sociodemographic factors associated with self-reported antidepressant non-response in UKB. Odds ratios, 95% confidence intervals significance, controlling for all other factors: SSRI (blue; N = 17,789), citalopram (pink; N = 7,636), fluoxetine (green; N = 7,750), paroxetine (purple; N = 2,119) and sertraline (yellow; N = 5,352). Ethnic background has been excluded from this figure because of wide confidence intervals (see Supplementary materials 2, Table S1).

### Factors associated with treatment response

#### Sociodemographic factors

In univariable analysis, the strongest factor (largest effect size and lowest p-value) associated with SSRI non-response across all phenotypes was self-medication with drugs and alcohol, followed by younger age, male gender, higher neighborhood deprivation, current smoking, lower income (<£18K), previous drinking, and mixed ethnicity (Supplementary materials2, TableS1).

For the composite-SSRI outcome, 17,789 participants had complete data across included predictors. Multivariable logistic regression showed self-medication, age, gender, and income remained significant predictors of non-response after multiple testing adjustment (Figure2, Supplementary materials2, TableS2). Specifically, self-medicating was associated with higher odds of reporting that SSRIs did not improve symptoms (OR=1.56, 95%CI=1.41-1.72, *p*=2.08×10^−19^) - i.e. those who self-medicated were more likely to report an SSRI did not help them improve (“feel better”) compared to those who did not self-medicate. Compared to older individuals, younger age was associated with an increased likelihood of SSRI non-response (OR=0.87, 95%CI=0.84-0.91, *p*=2.49 ×10^−11^), with the odds of non-response decreasing by 13% for each additional year. Male gender (OR=1.26, 95%CI=1.16-1.37, *p*=3.18×10^−08^) and lower income (£18K) relative to the median income (£30K-£52K) (OR=1.34, 95%CI=1.19-1.50, *p*=6.17×10^−07^) were also linked to non-response.

Neighborhood deprivation, smoking status, drinking status, and ethnicity were nominally significant but did not meet the multiple testing threshold (p<0.006). These patterns were consistent in the conservative-SSRI outcome (N=16,540), where TDI was also associated with non-response (OR=1.07, 95%CI=1.03-1.12, *p*=1.66×10^−03^). Sex-stratified analyses showed non-response was associated with self-medication, younger age, and lower income in females (N=13,049), and with self-medication and lower income in males (N=4,869) (Supplementary materials2, TableS2).

At the drug-specific level, treatment response associations mirrored the broader SSRI pattern, with some variation. Multivariable analysis showed that self-medication and lower income were consistently associated with all SSRI responses, while the impact of gender and smoking showed distinct associations depending on the drug. Ethnicity and educational attainment were not linked to non-response for any drug. Self-medication was the strongest predictor across SSRIs, particularly for citalopram (N=7,636) (OR=1.52, 95%CI=1.31-1.77, *p*=6.60×10^−08^) and sertraline (N=5,352) (OR=1.43, 95%CI=1.24-1.64, *p*=5.22×10^−07^). Lower income was another common predictor across drugs, including citalopram (OR=1.33, 95%CI=1.10-1.61, *p*=3.28×10^−03^) and sertraline (OR=1.54, 95%CI=1.26-1.89, *p*=2.99×10^−05^). Younger age was significantly associated with non-response to citalopram, paroxetine, and sertraline. The effect of sex differed between drugs; male gender was a stronger predictor for fluoxetine non-response (OR=1.53, 95%CI=1.36-1.73, *p*=5.64×10^−12^) compared to other SSRIs. Unique associations included an association between current smoking and citalopram non-response among males (OR=1.47, 95%CI= 1.14-1.91, *p*=3.03×10^−03^).

#### Clinical factors

In univariable analysis, clinical characteristics associated with non-response showed consistent patterns across overall SSRI and drug-specific outcomes (Supplementary materials2, TableS3). Non-response was associated with a later age at first episode, multiple episodes, worst episode duration exceeding two years, lack of mood brightening during the worst episode, difficulty coping with rejection, feelings of heavy limbs, worthlessness, and thoughts of death.

In multivariable analyses (Figure3, Supplementary materials2, TableS4), for the composite-SSRI outcome (N=9,237), a worst episode lasting more than two years (OR=1.97, 95%CI=1.68-2.31, *p*=1.25×10^−16^) and no brightening of mood in response to positive events during worst episode of depression (OR=1.33, 95%CI=1.19-1.50, *p*=1.34×10^−06^) were significantly associated with SSRI non-response. Similar patterns were observed in the conservative-SSRI model. At the drug-specific level, a duration over two years persisted as the strongest association with non-response across all drugs (citalopram: OR=2.13, 95%CI=1.65-2.75, *p*=8.22×10^−09^; fluoxetine: OR=1.64, 95%CI=1.2-2.09, *p*=6.48×10^−05^; paroxetine: OR=2.64, 95%CI=1.67-4.17, *p*=3.48×10^−05^; sertraline: OR=1.94, 95%CI=1.46-2.56, *p*=4.16×10^−06^). Lack of mood brightening was also associated with non-response to citalopram (OR=1.37, 95%CI=1.14-1.66, *p*=1.01×10^−03^) and fluoxetine (OR=1.39, 95%CI=1.16-1.67, *p*=3.18×10^−04^). The association between conservative-SSRI and brightening of mood did not persist when using a strict multiple testing threshold (p<0.0006).

**Figure 3:**
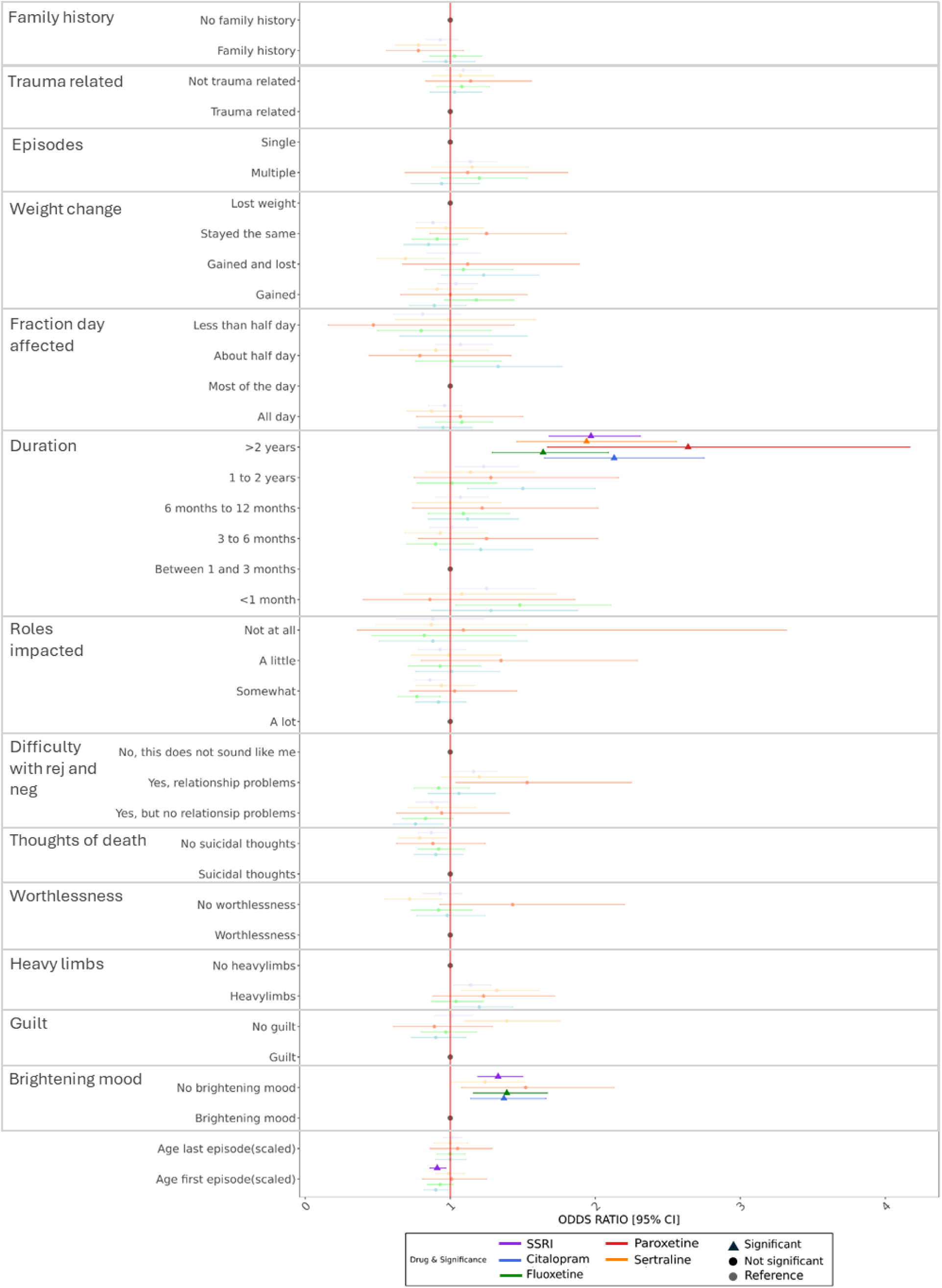
Clinical factors associated with self-reported antidepressant non-response in UKB. Odds ratios, 95% confidence intervals significance, controlling for all other factors: SSRI (blue; N = 9,237), citalopram (pink; N = 4072), fluoxetine (green; N = 3,917), paroxetine (purple; N = 1,099) and sertraline (yellow; N = 2,893). Analyses have been restricted to those of white ethnic background and adjusted for age and sex.

#### Genetic factors

##### CYP2C19 metabolizer status

Our study included 18,992 unrelated individuals of European ancestry with available CYP2C19 genotype and phenotype data. The largest group was CYP2C19 (7,507, 39.5%), followed by RM (5,155, 27.1%) and IM (4,943, 26.0%). As expected, PM (443, 2.3%) and UM (930, 4.9%) were less common, with 0.1% having indeterminate status. Similar proportions were observed at the drug-specific level (Supplementary materials1, Table S11).

Association analyses between self-reported antidepressant response and CYP2C19 metabolizer status shows, among SSRI users, PM had nominally significant higher likelihood of non-response compared to NM (OR=1.31, 95%CI=1.05-1.65, *p*=1.77×10^−02^); this was mirrored in the conservative approach but with increased significance (OR=1.41, 95%CI=1.09-1.83, p=8.57×10^−03^). A similar association was observed for fluoxetine (OR=1.42, 95%CI=1.02-1.97, *p*=3.50×10^−02^) (Figure4, Supplementary materials2, TableS5).

**Figure 4:**
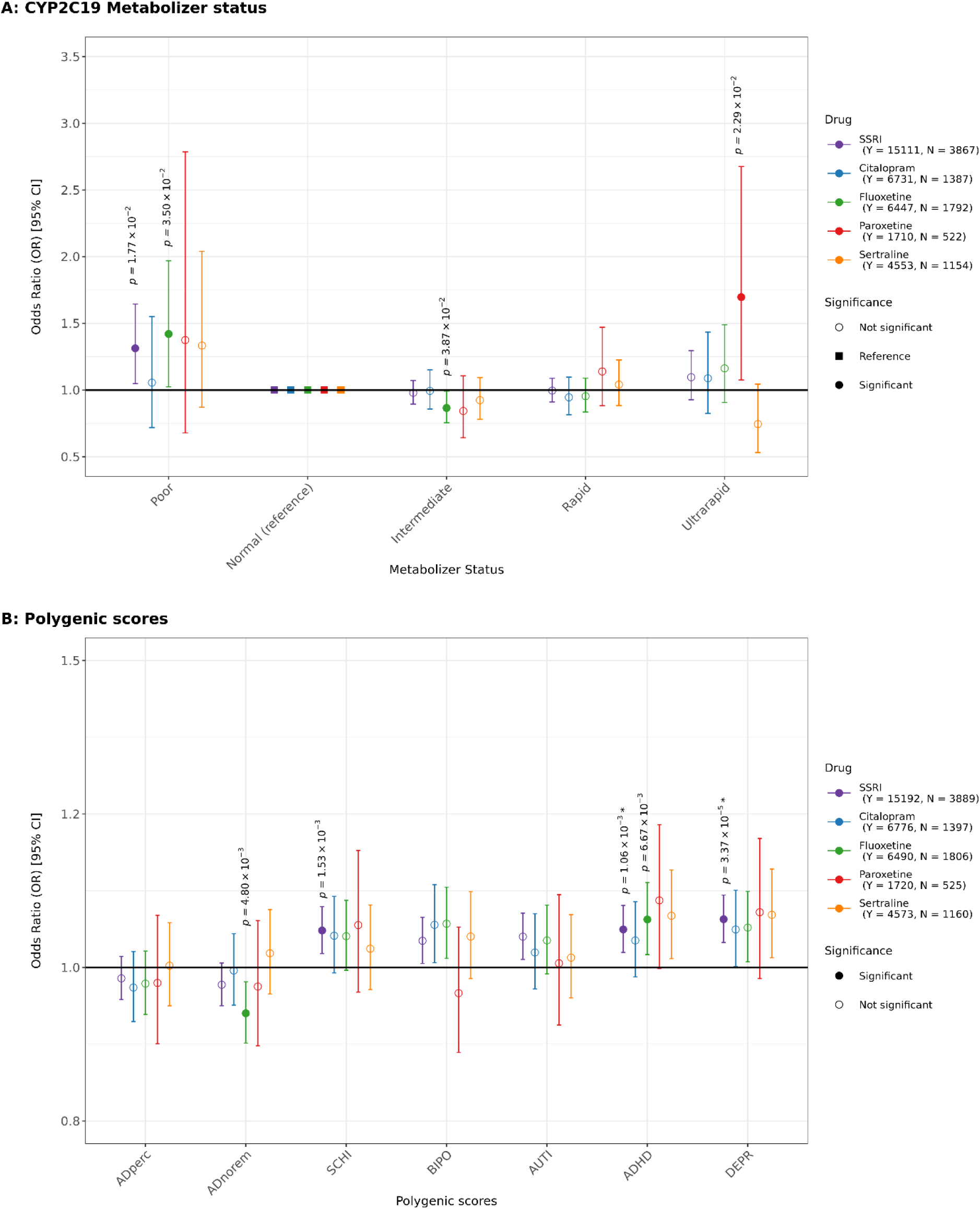
Genetic predictors of self-reported antidepressant non-response in UKB. A: **Inferred CYP2C19 metabolizer status associated with self-reported antidepressant non-response in UKB**. Forest plots depict odds ratios and 95% confidence intervals for the association between metabolizer category (compared to normal) and treatment non-response (blue markers) for SSRIs and specific SSRIs (Citalopram, Fluoxetine, Paroxetine, Sertraline). Inferred metabolizer status levels are Poor, Normal (reference), Intermediate, Rapid, and Ultrarapid. Multiple testing correction is not required as a single predictor. Significance: p<0.05; *p indicates significant persisted after stringent correction for multiple testing (P< 0.008). **B. Psychiatric and antidepressant response PGS associated with self-reported antidepressant non-response in UKB**. The association analyses between SSRIs and specific SSRIs (Citalopram, Fluoxetine, Paroxetine, Sertraline) and self-reported antidepressant outcomes and various mental health condition and treatment polygenic scores (PGS). PGS include DEPR: Depression, ADHD: Attention Deficit Hyperactivity Disorder, AUTI: Autism, BIPO: Bipolar Disorder, SCHI: Schizophrenia. ADperc: Percentage improvement, ADnorem: AD non-remission. *indicates significance persisted under strict multiple testing correction (P<0.007).

Drug-specific associations showed lower odds of self-reported non-response among IM using fluoxetine (OR=0.87, 95%CI=0.76 =0.99, *p*=3.87×10^−02^) and higher odds of non-response among UM taking paroxetine (OR=1.70, 95%CI=1.08-2.68, *p*=2.29×10^−02^). No significant associations were noted among citalopram and sertraline users. None of the identified associations survived multiple testing correction that accounts for the six response outcomes (*p*=0.008).

##### Polygenic scores

We tested for the association between self-reported antidepressant response and PGS for five psychiatric disorders and two antidepressant response measures (Figure4, Supplementary materials2, TableS6). For psychiatric traits, composite-SSRI (“SSRI” in Figure 5) non-response was significantly associated with PGS for depression (OR=1.08, 95%CI=1.04-1.12, *p*=3.37×10^−05^), attention deficit hyperactivity disorder (ADHD) (OR=1.06, 95%CI=1.02-1.10, *p*=1.06×10^−03^) and schizophrenia (OR=1.06, 95%CI=1.02-1.10, *p*=1.53×10^−03^). In each case, higher genetic liability to the disorder (higher PGS) corresponded with a lack of response to SSRIs (6-8% increase in the odds of non-response per sd increase of PGS). The phenotypic variance explained (*R^2^*) for each of these PGS with composite-SSRI non-response was 0.16%, 0.11% and 0.1% respectively (Supplementary materials1, Figure S4)

In the drug-specific analysis, the only significant finding after multiple testing correction was for fluoxetine and the ADHD PGS (*R^2^*=0.22%, OR=1.08, 95%CI=1.02-1.14, *p*= 6.67×10^− 03^). Other disorders showed nominal significance (e.g. depression and bipolar PGS with citalopram and fluoxetine; and depression and ADHD PGS with sertraline) (Figure5).

For the antidepressant response PGS (non-remission (ADnorem) and percentage improvement (ADperc)), the only significant finding after multiple testing correction was a negative association between non-response to fluoxetine and the non-remission PGS (*R^2^*=0.235, OR=0.93, 95%CI=0.88-0.98, *p*=4.80×10^−03^). However, this association did not persist with a stringent correction (p<0.001).

## DISCUSSION

Antidepressants, particularly SSRIs, are widely prescribed, yet remission rates remain low, and few risk factors guide personalized prescribing. A major challenge in identifying factors to predict treatment is the absence of response measures that are scalable across large sample sizes. Several large studies have applied patient self-report questionnaires on response, asking a single, simple question about whether an antidepressant has ‘helped’. In this paper we assessed the UKB’s antidepressant self-report data to identify clinical, sociodemographic, and genetic factors associated with a lack of response to SSRIs, aiming to demonstrate that this self-reported outcome is a valid measure of antidepressant response.

In UKB, 75% of participants reported a positive SSRI response, substantially higher than the 35% remission rate noted in trials (Rush et al., 2004). This discrepancy may reflect the question wording (“Has <drugname> helped you feel better” with possible responses of “Yes, at least a little”, and “No”), which is not synonymous with remission. This is a lower threshold than the criteria used for remission in clinical trials, where depression symptoms must fall below the diagnostic threshold for depression (Stone et al., 2022).

For self-reported antidepressant response to be a valuable phenotype, a deeper understanding of factors associated with a negative response is needed. Our analysis revealed significant associations with sociodemographic, clinical and genetic variables, with consistent direction and effect sizes across SSRIs. Non-response to SSRIs was associated with being younger, male, having a lower income and self-medicating with drugs and alcohol. These factors largely align with clinical trial predictors while also highlighting real-world influences like substance use.

For example, males exhibited lower response rates, consistent with some clinical studies (Gibiino, Marsano, & Serretti, 2014; Serretti, Gibiino, & Drago, 2011; Trivedi et al., 2006), although no difference was found in iSPOT-D (Saveanu et al., 2015) or a meta-analysis of Randomised Control Trials (RCTs) (Cuijpers et al., 2014). Younger age at UKB recruitment was associated with better SSRI response; this aligns with data suggesting depression is more chronic and antidepressants are less effective in older patients (Haigh, Bogucki, Sigmon, & Blazer, 2018; Strawn et al., 2023). However, no information on age at antidepressant use is available in UKB. Lower income has been associated with reduced treatment response in a recent RCT (Mills et al., 2022), and in STAR*D, where reduced adherence and shorter treatment duration was found in lower income groups, even when controlled for treatment access and level of care (Jakubovski & Bloch, 2014).

Clinical associations of poor SSRI response in UKB were persistent low mood and long depressive episodes (≥2 years), consistent with findings that greater severity (Rush et al., 2004) and melancholic depression hinder recovery and reduce perceived antidepressant efficacy (Valerio, Szmulewicz, & Martino, 2018). Hieronymus et al. emphasize targeting low mood to improve MDD treatment outcomes; their SSRI RCT reanalysis found higher rates of efficacy when focusing on mood improvements (where 91% of participants showed efficacy) than with HDRS-17 scores (efficacy in 44%) (Hieronymus, Emilsson, Nilsson, & Eriksson, 2016). However, unlike clinical trials where greater severity predicts better response to antidepressants than placebo, in our study, proxy measures of higher severity — determined by long-duration episodes (≥2 years) and higher genetic liability for MDD — were correlated with non-response.

Our study provides insights into the genetic basis for SSRI response. Higher genetic liability for depression, schizophrenia, and attention deficit/hyperactivity disorder (ADHD) was associated with poorer self-reported antidepressant response, consistent with previous research (Pain et al., 2022). The association between higher PGS and treatment non-response is consistent with clinical studies that link higher depression PGS to increased side effects, such as dizziness and reduced sexual desire, which may impact treatment effectiveness by hindering effective dosage maintenance (Campos et al., 2021). Higher schizophrenia PGS in non-responders may reflect elevated rates of progression to schizophrenia and bipolar disorder post-MDD diagnosis, as seen in iPSYCH (Als et al., 2023). The link between genetic liability for ADHD in UKB may reflect missed diagnosis in this older age-group. A recent study reported that 28% of adults referred for mood and anxiety assessments had undiagnosed ADHD, with the number of prior SSRI prescriptions being a significant predictor (Sternat, Mohammed, & Furtado, 2016).

The PGS for non-remission from clinical trials was – counter-intuitively – associated with a positive response to SSRIs in our studies (Pain et al., 2022). This discrepancy may be partly due to individuals with a higher genetic predisposition for non-remission experiencing persistent symptoms that require prolonged treatment. In such cases, antidepressant efficacy may be attributable to providing day-to-day symptom management rather than remission or coincide with the natural progression of an episode. The perception of antidepressant efficacy based on improved daily functioning aligns with findings that patients prioritize a return to normal functioning as much as the absence of symptoms (Zimmerman et al., 2006). Additionally, unlike the 8–12-week assessment span of clinical trials, the UKB questionnaire uses a retrospective measure based on the full length of an SSRI treatment period, suggesting while self-reported response may not adequately gauge ‘remission’ status, it could reflect improvements in personal functioning.

CYP2C19 metabolizer status proportions in our study were highly concordant to other studies in European populations (Campos et al., 2022; Ionova et al., 2020). The positive association between PM status and SSRI non-response is as expected given reduced CYP2C19 enzyme among PM slows drug metabolism, leading to suboptimal drug levels, prolonged exposure, increased side effects, and inadequate therapeutic response (Campos et al., 2022; Ionova et al., 2020). At the drug-specific level, fluoxetine also showed this association and additionally demonstrated an increased response among IM.

Non-response to paroxetine in UM suggests insufficient drug levels to achieve therapeutic effects, necessitating higher doses (Li et al., 2024; Wong et al., 2023). No effect of metabolizer status was observed among those taking citalopram and sertraline which aligns with previous work assessing the impact of CYP2C19 variation on drug tolerability (Campos et al., 2022); where the authors postulate that the wide therapeutic windows of these drugs may limit the ability of CYP2C19 polymorphisms to impact treatment response. We note that CYP2C19 is not the sole metabolizing enzyme: although primarily responsible for metabolizing citalopram, it has a lesser role for sertraline, fluoxetine, and paroxetine (Li et al., 2024; Obach, Cox, & Tremaine, 2005; Yuce-Artun et al., 2016).

Mental health research increasingly recognizes the importance of patient perspectives and priorities, but there is a paucity of studies on patients’ views on antidepressant use and response. The Wellcome Trust has prioritized patient and public involvement (PPI) in all their funded Mental Health studies (Wellcome Trust, n.d.), and our Antidepressant Medications: Biology, Exposure and Response (AMBER) research program has ongoing research to understand the priorities of those taking antidepressants.

Self-report questionnaires from the UKB provide novel and rich data to better understand patient perspectives, which can, in turn, help more accurately measure outcomes and tailor management pathways. Despite its potential utility, additional measures, such as a refined scale that captures multiple types of positive responses, could further enhance our understanding of what constitutes meaningful recovery for patients.

This study has many strengths in its patient-centered approach, providing valuable information on antidepressant treatment from the patient’s perspective. This easily collected phenotype is available in large numbers in the UKB, the Australian Genetics of Depression study (AGDS, (Byrne et al., 2020)), and Genetic Links to Anxiety and Depression (GLAD) (Davies et al., 2019; Koch et al., 2024). Our study shows good alignment of sociodemographic and clinical risk factors between the UKB self-report response and clinical trial research in antidepressants, suggesting both measures pick up common information on response and non-response. This provides an important source of information to expand sample sizes for research into the genetic underpinnings of antidepressant response. Differences between clinician-rated and self-rated measures suggest patients have unique perspectives on treatment outcomes, which are critical to a complete assessment of depression (Campos et al., 2022; Ramanuj, Ferenchick, & Pincus, 2019; Uher et al., 2012; Zimmerman et al., 2006).

While our study offers valuable insights into antidepressant response, it has several limitations. The analyses were based on UKB, which has better-than-average socioeconomic circumstances than the general population, potentially limiting the generalizability of the findings and underestimating the impact of low income. The reliance on retrospective self-reports is subject to recall bias and inaccuracies - participants may either underreport or overreport their past symptoms and healthcare experiences. Sociodemographic variables were reported at the time of the baseline UKB questionnaire, not during the participant’s worst depressive episode, complicating factor assessment. Further limitations of this analysis include its focus on those who have tried SSRIs, potentially resulting in a dataset that may not fully capture the diversity of antidepressant usage amongst the UK population. Moreover, the inability to determine the timing of antidepressant exposure and whether the drugs were used independently or concurrently, or whether they were prescribed for different episodes or had overlapping use, complicates the interpretation of treatment patterns and outcomes. We are unable to dissect natural disorder course from antidepressant response, and some patients responding that the drug helped them may have had similar recovery trajectory without an antidepressant.

## CONCLUSION

Self-reported antidepressant response outcome in UK Biobank is influenced by sociodemographic, clinical characteristics and common genetic variation, similar to clinical response measures. Despite the higher frequency of positive response outcomes compared to clinical trials, these retrospective self-report outcomes replicate known associations with current antidepressant treatment outcomes. This suggests that self-reported outcomes, while measuring a positive response that is not equivalent to remission, capture meaningful aspects of antidepressant effectiveness, particularly from the patient’s perspective.

## Supplementary material

The supplementary materials for this article include Supplementary materials 1 and Supplementary materials 2.

## Data availability statement

Code for GenoPred Pipeline is available from the GenoPred GitHub Repository (https://opain.github.io/GenoPred/). Summary statistics from the PGC are available online (https://pgc.unc.edu/for-researchers/download-results/). Data from the UK Biobank (https://www.ukbiobank.ac.uk) are available to bona fide researchers upon application.

## Acknowledgements

We gratefully acknowledge the time and attention of the participants who generously provided data and donated their samples to the UK Biobank. UKB analysis conducted under project 82087. This work made use of the King’s Computational Research, Engineering and Technology Environment (CREATE) provided by e-Research King’s College London (https://docs.er.kcl.ac.uk/). This publication is the work of the authors and CML will serve as guarantor for the contents of this paper.

## Declarations

CML sits on the Myriad Neuroscience Scientific Advisory Board and is a Key Opinion Leader for UCB Pharma. OP provides consultancy services for UCB pharma company. The remaining authors declare that there are no competing interests.

The funders had no role in the study’s design; in the collection, analyses, or interpretation of data; in the writing of the manuscript, or in the decision to publish the results.

The authors assert that all procedures contributing to this work comply with the ethical standards of the relevant national and institutional committees on human experimentation and with the Helsinki Declaration of 1975, as revised in 2008.

## REFERENCES

Abdoli, N., Salari, N., Darvishi, N., Jafarpour, S., Solaymani, M., Mohammadi, M., & Shohaimi, S. (2022). The global prevalence of major depressive disorder (MDD) among the elderly: A systematic review and meta-analysis. Neuroscience and Biobehavioral Reviews, 132, 1067–1073. 10.1016/j.neubiorev.2021.10.041

Allen, N. E., Sudlow, C., Peakman, T., & Collins, R. (2014). UK Biobank Data: Come and Get It. Science Translational Medicine, 6(224), 224ed4–224ed4. 10.1126/scitranslmed.3008601

Als, T. D., Kurki, M. I., Grove, J., Voloudakis, G., Therrien, K., Tasanko, E., … Børglum, A. D. (2023). Depression pathophysiology, risk prediction of recurrence and comorbid psychiatric disorders using genome-wide analyses. Nature Medicine, 29(7), 1832–1844. 10.1038/s41591-023-02352-1

Bauer, M., Severus, E., Möller, H.-J., & Young, A. H. (2017). Pharmacological treatment of unipolar depressive disorders: summary of WFSBP guidelines. International Journal of Psychiatry in Clinical Practice, 21(3), 166–176. 10.1080/13651501.2017.1306082

Byrne, E. M., Kirk, K. M., Medland, S. E., McGrath, J. J., Colodro-Conde, L., Parker, R., … Martin, N. G. (2020). Cohort profile: the Australian genetics of depression study. BMJ Open, 10(5), e032580. 10.1136/bmjopen-2019-032580

Campos, A. I., Byrne, E. M., Mitchell, B. L., Wray, N. R., Lind, P. A., Licinio, J., … Rentería, M. E. (2022). Impact of CYP2C19 metaboliser status on SSRI response: a retrospective study of 9500 participants of the Australian Genetics of Depression Study. The Pharmacogenomics Journal, 22(2), 130–135. 10.1038/s41397-022-00267-7

Campos, A. I., Mulcahy, A., Thorp, J. G., Wray, N. R., Byrne, E. M., Lind, P. A., … Rentería, M. E. (2021). Understanding genetic risk factors for common side effects of antidepressant medications. Communications Medicine, 1, 45. 10.1038/s43856-021-00046-8

Cipriani, A., Furukawa, T. A., Salanti, G., Chaimani, A., Atkinson, L. Z., Ogawa, Y., … Geddes, J. R. (2018). Comparative Efficacy and Acceptability of 21 Antidepressant Drugs for the Acute Treatment of Adults With Major Depressive Disorder: A Systematic Review and Network Meta-Analysis. Focus (American Psychiatric Publishing), 16(4), 420–429. 10.1176/appi.focus.16407

Cleare, A., Pariante, C. M., Young, A. H., Anderson, I. M., Christmas, D., Cowen, P. J., … Uher, R. (2015). Evidence-based guidelines for treating depressive disorders with antidepressants: A revision of the 2008 British Association for Psychopharmacology guidelines. Journal of Psychopharmacology, 29(5), 459–525. 10.1177/0269881115581093

Cui, L., Li, S., Wang, S., Wu, X., Liu, Y., Yu, W., … Li, B. (2024). Major depressive disorder: hypothesis, mechanism, prevention and treatment. Signal Transduction and Targeted Therapy 2024 9:1, 9(1), 1–32. 10.1038/s41392-024-01738-y

Cuijpers, P., Weitz, E., Twisk, J., Kuehner, C., Cristea, I., David, D., … Hollon, S. D. (2014). Gender as predictor and moderator of outcome in cognitive behavior therapy and pharmacotherapy for adult depression: an “individual patient data” meta-analysis. Depression and Anxiety, 31(11), 941–951. 10.1002/da.22328

Davies, M. R., Kalsi, G., Armour, C., Jones, I. R., McIntosh, A. M., Smith, D. J., … Breen, G. (2019). The Genetic Links to Anxiety and Depression (GLAD) Study: Online recruitment into the largest recontactable study of depression and anxiety. Behaviour Research and Therapy, 123, 103503. 10.1016/j.brat.2019.103503

Demontis, D., Walters, R. K., Martin, J., Mattheisen, M., Als, T. D., Agerbo, E., … Neale, B. M. (2019). Discovery of the first genome-wide significant risk loci for attention deficit/hyperactivity disorder. Nature Genetics, 51(1), 63–75. 10.1038/s41588-018-0269-7

Fanelli, G., Benedetti, F., Kasper, S., Zohar, J., Souery, D., Montgomery, S., … Fabbri, C. (2021). Higher polygenic risk scores for schizophrenia may be suggestive of treatment non-response in major depressive disorder. Progress in Neuro-Psychopharmacology & Biological Psychiatry, 108, 110170. 10.1016/j.pnpbp.2020.110170

Fried, E. I., & Nesse, R. M. (2015). Depression is not a consistent syndrome: An investigation of unique symptom patterns in the STAR*D study. Journal of Affective Disorders, 172, 96–102. 10.1016/j.jad.2014.10.010

García-González, J., Tansey, K. E., Hauser, J., Henigsberg, N., Maier, W., Mors, O., … Fabbri, C. (2017). Pharmacogenetics of antidepressant response: A polygenic approach. Progress in Neuro-Psychopharmacology & Biological Psychiatry, 75, 128–134. 10.1016/j.pnpbp.2017.01.011

GBD 2019 Mental Disorders Collaborators. (2022). Global, regional, and national burden of 12 mental disorders in 204 countries and territories, 1990-2019: a systematic analysis for the Global Burden of Disease Study 2019. The Lancet Psychiatry, 9(2), 137–150. 10.1016/S2215-0366(21)00395-3

Gibiino, S., Marsano, A., & Serretti, A. (2014). Specificity profile of venlafaxine and sertraline in major depression: metaregression of double-blind, randomized clinical trials. The International Journal of Neuropsychopharmacology, 17(1), 1–8. 10.1017/S1461145713000746

Grove, J., Ripke, S., Als, T. D., Mattheisen, M., Walters, R. K., Won, H., … Børglum, A. D. (2019). Identification of common genetic risk variants for autism spectrum disorder. Nature Genetics, 51(3), 431–444. 10.1038/s41588-019-0344-8

Haigh, E. A. P., Bogucki, O. E., Sigmon, S. T., & Blazer, D. G. (2018). Depression Among Older Adults: A 20-Year Update on Five Common Myths and Misconceptions. The American Journal of Geriatric Psychiatry : Official Journal of the American Association for Geriatric Psychiatry, 26(1), 107–122. 10.1016/j.jagp.2017.06.011

Hieronymus, F., Emilsson, J. F., Nilsson, S., & Eriksson, E. (2016). Consistent superiority of selective serotonin reuptake inhibitors over placebo in reducing depressed mood in patients with major depression. Molecular Psychiatry, 21(4), 523–530. 10.1038/mp.2015.53

Iacobucci, G. (2019). NHS prescribed record number of antidepressants last year. BMJ (Clinical Research Ed.), 364, l1508. 10.1136/BMJ.L1508

Ionova, Y., Ashenhurst, J., Zhan, J., Nhan, H., Kosinski, C., Tamraz, B., & Chubb, A. (2020). CYP2C19 Allele Frequencies in Over 2.2 Million Direct-to-Consumer Genetics Research Participants and the Potential Implication for Prescriptions in a Large Health System. Clinical and Translational Science, 13(6), 1298–1306. 10.1111/cts.12830

Ising, M., Lucae, S., Binder, E. B., Bettecken, T., Uhr, M., Ripke, S., … Müller-Myhsok, B. (2009). A Genomewide Association Study Points to Multiple Loci That Predict Antidepressant Drug Treatment Outcome in Depression. Archives of General Psychiatry, 66(9), 966–975. 10.1001/archgenpsychiatry.2009.95

Jakubovski, E., & Bloch, M. H. (2014). Prognostic subgroups for citalopram response in the STAR*D trial. The Journal of Clinical Psychiatry, 75(7), 738–747. 10.4088/JCP.13m08727

Ji, Y., Biernacka, J. M., Hebbring, S., Chai, Y., Jenkins, G. D., Batzler, A., … Mrazek, D. A. (2013). Pharmacogenomics of selective serotonin reuptake inhibitor treatment for major depressive disorder: genome-wide associations and functional genomics. The Pharmacogenomics Journal, 13(5), 456–463. 10.1038/tpj.2012.32

Karrouri, R., Hammani, Z., Benjelloun, R., & Otheman, Y. (2021). Major depressive disorder: Validated treatments and future challenges. World Journal of Clinical Cases, 9(31), 9350–9367. 10.12998/wjcc.v9.i31.9350

Kessler, R. C., van Loo, H. M., Wardenaar, K. J., Bossarte, R. M., Brenner, L. A., Ebert, D. D., … Zaslavsky, A. M. (2017). Using patient self-reports to study heterogeneity of treatment effects in major depressive disorder. Epidemiology and Psychiatric Sciences, 26(1), 22–36. DOI: 10.1017/S2045796016000020

Khan, A., Brodhead, A. E., Schwartz, K. A., Kolts, R. L., & Brown, W. A. (2005). Sex Differences in Antidepressant Response in Recent Antidepressant Clinical Trials. Journal of Clinical Psychopharmacology, 25(4). Retrieved from https://journals.lww.com/psychopharmacology/fulltext/2005/08000/sex_differences_in_antidepressant_response_in.5.aspx

Koch, E., Jürgenson, T., Einarsson, G., Mitchell, B., Harder, A., Garc\’\ia-Mar\’\in, L. M., … O\textquoterightConnell, K. S. (2024). Genome-wide meta-analyses of non-response to antidepressants identify novel loci and potential drugs. MedRxiv. 10.1101/2024.07.13.24310361

Li, D., Pain, O., Chiara, F., Wong, W. L. E., Lo, C. W. H., Ripke, S., … the GSRD Consortium, the M. D. D. W. G. of the P. G. C. (2024). Metabolic activity of CYP2C19 and CYP2D6 on antidepressant response from 13 clinical studies using genotype imputation: a meta-analysis. Translational Psychiatry, 14(1), 296. 10.1038/s41398-024-02981-1

McInnes, G., Lavertu, A., Sangkuhl, K., Klein, T. E., Whirl-Carrillo, M., & Altman, R. B. (2021). Pharmacogenetics at Scale: An Analysis of the UK Biobank. Clinical Pharmacology and Therapeutics, 109(6), 1528–1537. 10.1002/cpt.2122

Mills, J. A., Suresh, V., Chang, L., Mayes, T., Croarkin, P. E., Trivedi, M. H., & Strawn, J. R. (2022). Socioeconomic Predictors of Treatment Outcomes Among Adults With Major Depressive Disorder. Psychiatric Services (Washington, D.C.), 73(9), 965–969. 10.1176/appi.ps.202100559

Ni, G., Zeng, J., Revez, J. A., Wang, Y., Zheng, Z., Ge, T., … Wray, N. R. (2021). A Comparison of Ten Polygenic Score Methods for Psychiatric Disorders Applied Across Multiple Cohorts. Biological Psychiatry, 90(9), 611–620. 10.1016/j.biopsych.2021.04.018

NICE. Depression in adults: Recognition and management., (2009).

Obach, R. S., Cox, L. M., & Tremaine, L. M. (2005). Sertraline is metabolized by multiple cytochrome P450 enzymes, monoamine oxidases, and glucuronyl transferases in human: an in vitro study. Drug Metabolism and Disposition, 33(2), 262 LP – 270. 10.1124/dmd.104.002428

Otte, C., Gold, S. M., Penninx, B. W., Pariante, C. M., Etkin, A., Fava, M., … Schatzberg, A. F. (2016). Major depressive disorder. Nature Reviews Disease Primers 2016 2:1, 2(1), 1–20. 10.1038/nrdp.2016.65

Pain, O., Al-Chalabi, A., & Lewis, C. (2024). The GenoPred Pipeline: A Comprehensive and Scalable Pipeline for Polygenic Scoring. 10.1101/2024.06.12.24308843

Pain, O., Hodgson, K., Trubetskoy, V., Ripke, S., Marshe, V. S., Adams, M. J., … Lewis, C. M. (2022). Identifying the Common Genetic Basis of Antidepressant Response. Biological Psychiatry Global Open Science, 2(2), 115–126. 10.1016/j.bpsgos.2021.07.008

Pardiñas, A. F., Holmans, P., Pocklington, A. J., Escott-Price, V., Ripke, S., Carrera, N., … Walters, J. T. R. (2018). Common schizophrenia alleles are enriched in mutation-intolerant genes and in regions under strong background selection. Nature Genetics, 50(3), 381–389. 10.1038/s41588-018-0059-2

Perna, G., Alciati, A., Daccò, S., Grassi, M., & Caldirola, D. (2020). Personalized Psychiatry and Depression: The Role of Sociodemographic and Clinical Variables. Psychiatry Investigation, 17(3), 193–206. 10.30773/pi.2019.0289

R Core Team. (2021). R: A language and environment for statistical computing. Vienna, Austria: R Foundation for Statistical Computing. Retrieved from https://www.r-project.org/

Ramanuj, P., Ferenchick, E. K., & Pincus, H. A. (2019). Depression in primary care: part 2-management. BMJ (Clinical Research Ed.), 365, l835. 10.1136/bmj.l835

Rayner, C., Coleman, J. R. I., Purves, K. L., Carr, E., Cheesman, R., Davies, M. R., … Eley, T. C. (2021). Sociodemographic factors associated with treatment-seeking and treatment receipt: cross-sectional analysis of UK Biobank participants with lifetime generalised anxiety or major depressive disorder. BJPsych Open, 7(6), e216. DOI: 10.1192/bjo.2021.1012

Rush, A. J., Fava, M., Wisniewski, S. R., Lavori, P. W., Trivedi, M. H., Sackeim, H. A., … for the STAR*D Investigators Group. (2004). Sequenced treatment alternatives to relieve depression (STAR*D): rationale and design. Controlled Clinical Trials, 25(1), 119–142. 10.1016/S0197-2456(03)00112-0

Rush, A. J., Trivedi, M. H., Stewart, J. W., Nierenberg, A. A., Fava, M., Kurian, B. T., … Wisniewski, S. R. (2011). Combining Medications to Enhance Depression Outcomes (CO-MED): Acute and Long-Term Outcomes of a Single-Blind Randomized Study. American Journal of Psychiatry, 168(7), 689–701. 10.1176/appi.ajp.2011.10111645

Rush, A. J., Trivedi, M. H., Wisniewski, S. R., Nierenberg, A. A., Stewart, J. W., Warden, D., … Fava, M. (2006). Acute and longer-term outcomes in depressed outpatients requiring one or several treatment steps: a STAR*D report. The American Journal of Psychiatry, 163(11), 1905–1917. 10.1176/ajp.2006.163.11.1905

Sangkuhl, K., Whirl-Carrillo, M., Whaley, R. M., Woon, M., Lavertu, A., Altman, R. B., … Klein, T. E. (2020). Pharmacogenomics Clinical Annotation Tool (PharmCAT). Clinical Pharmacology & Therapeutics, 107(1), 203–210. 10.1002/cpt.1568

Saveanu, R., Etkin, A., Duchemin, A.-M., Goldstein-Piekarski, A., Gyurak, A., Debattista, C., … Williams, L. M. (2015). The international Study to Predict Optimized Treatment in Depression (iSPOT-D): outcomes from the acute phase of antidepressant treatment. Journal of Psychiatric Research, 61, 1–12. 10.1016/j.jpsychires.2014.12.018

Serretti, A., Gibiino, S., & Drago, A. (2011). Specificity profile of paroxetine in major depressive disorder: meta-regression of double-blind, randomized clinical trials. Journal of Affective Disorders, 132(1–2), 14–25. 10.1016/j.jad.2010.08.018

Souery, D., Serretti, A., Calati, R., Oswald, P., Massat, I., Konstantinidis, A., … Mendlewicz, J. (2011). Switching antidepressant class does not improve response or remission in treatment-resistant depression. Journal of Clinical Psychopharmacology, 31(4), 512–516. 10.1097/JCP.0b013e3182228619

Stahl, E. A., Breen, G., Forstner, A. J., McQuillin, A., Ripke, S., Trubetskoy, V., … Sklar, P. (2019). Genome-wide association study identifies 30 loci associated with bipolar disorder. Nature Genetics, 51(5), 793–803. 10.1038/s41588-019-0397-8

Sternat, M., Mohammed, T., & Furtado, T. (2016). SSRI treatment response may predict undetected attention deficit hyperactivity disorder in depressed patients. Anxiety and Depression Association of America (ADAA) Conference 2016, Abstract S2–10.

Stone, M. B., Yaseen, Z. S., Miller, B. J., Richardville, K., Kalaria, S. N., & Kirsch, I. (2022). Response to acute monotherapy for major depressive disorder in randomized, placebo controlled trials submitted to the US Food and Drug Administration: individual participant data analysis. BMJ, 378. 10.1136/bmj-2021-067606

Strawn, J. R., Mills, J. A., Suresh, V., Mayes, T., Gentry, M. T., Trivedi, M., & Croarkin, P. E. (2023). The impact of age on antidepressant response: A mega-analysis of individuals with major depressive disorder. Journal of Psychiatric Research, 159, 266–273. 10.1016/j.jpsychires.2023.01.043

Tansey, K. E., Guipponi, M., Hu, X., Domenici, E., Lewis, G., Malafosse, A., … Uher, R. (2013). Contribution of Common Genetic Variants to Antidepressant Response. Biological Psychiatry, 73(7), 679–682. 10.1016/j.biopsych.2012.10.030

Trivedi, M. H., Rush, A. J., Wisniewski, S. R., Nierenberg, A. A., Warden, D., Ritz, L., … Fava, M. (2006). Evaluation of Outcomes With Citalopram for Depression Using Measurement-Based Care in STAR*D: Implications for Clinical Practice. American Journal of Psychiatry, 163(1), 28–40. 10.1176/appi.ajp.163.1.28

Valerio, M. P., Szmulewicz, A. G., & Martino, D. J. (2018). A quantitative review on outcome-to-antidepressants in melancholic unipolar depression. Psychiatry Research, 265, 100–110. 10.1016/j.psychres.2018.03.088

Wellcome Trust. (n.d.). Embedding lived experience expertise in mental health research. Retrieved September 16, 2024, from https://wellcome.org/grant-funding/guidance/embedding-lived-experience-expertise-mental-health-research

Wong, W. L. E., Fabbri, C., Laplace, B., Li, D., van Westrhenen, R., Lewis, C. M., … Young, A. H. (2023). The Effects of CYP2C19 Genotype on Proxies of SSRI Antidepressant Response in the UK Biobank. Pharmaceuticals (Basel, Switzerland), 16(9). 10.3390/ph16091277

World Health Organization. (1993). Composite international Diagnostic Interview – Version 1.1. Geneva: WHO. 10.1163/_q3_SIM_00374

Wray, N. R., Ripke, S., Mattheisen, M., Trzaskowski, M., Byrne, E. M., Abdellaoui, A., … Sullivan, P. F. (2018). Genome-wide association analyses identify 44 risk variants and refine the genetic architecture of major depression. Nature Genetics, 50(5), 668–681. 10.1038/s41588-018-0090-3

Yuce-Artun, N., Baskak, B., Ozel-Kizil, E. T., Ozdemir, H., Uckun, Z., Devrimci-Ozguven, H., & Suzen, H. S. (2016). Influence of CYP2B6 and CYP2C19 polymorphisms on sertraline metabolism in major depression patients. International Journal of Clinical Pharmacy, 38(2), 388–394. 10.1007/s11096-016-0259-8

Zimmerman, M., McGlinchey, J. B., Posternak, M. A., Friedman, M., Attiullah, N., & Boerescu, D. (2006). How should remission from depression be defined? The depressed patient’s perspective. The American Journal of Psychiatry, 163(1), 148–150. 10.1176/appi.ajp.163.1.148

