## Supplementary Materials 1 for "Sociodemographic, clinical, and genetic factors associated with self-reported antidepressant response outcomes in the UK Biobank"

### Phenotype identification

#### Table S1: MHQ2 questionnaire UKB data fields used to identify SSRI users

| UKB field ID | Item | Answers |
|---|---|---|
| 29011 | Have you ever had a time in your life when you felt sad, blue, or depressed for two weeks or more in a row? | Yes<br>No<br>Do not know<br>Prefer not to answer |
| 29012 | Have you ever had a time in your life lasting two weeks or more when you lost interest in most things like hobbies, work, or activities that usually give you pleasure? | Yes<br>No<br>Do not know<br>Prefer not to answer |
| 29038 | Have you ever tried the following for these problems? | Unprescribed medication (more than once)<br>Medication prescribed to participant (for at least two weeks)<br>Drugs or alcohol (more than once)<br>None of the options listed<br>Prefer not to answer |
| 29039 | Have you ever tried any of the following medications for at least two weeks? | Citalopram (sometimes called Cipramil)<br>Fluoxetine (Prozac or Oxactin)<br>Sertraline (Lustral)<br>Paroxetine (Seroxat)<br>Amitriptyline (Elavil)<br>Dosulepin (Prothiaden)<br>Other antidepressant(s)<br>Do not know<br>Prefer not to answer |
| 29040 | Has citalopram helped you to feel better? | Yes, at least a little<br>No<br>Do not know<br>Prefer not to answer |
| 29041 | Has fluoxetine helped you to feel better? | |
| 29042 | Has sertraline helped you to feel better? | |
| 29043 | Has paroxetine helped you to feel better? | |

**UKB – UK Biobank*

### Study Participants

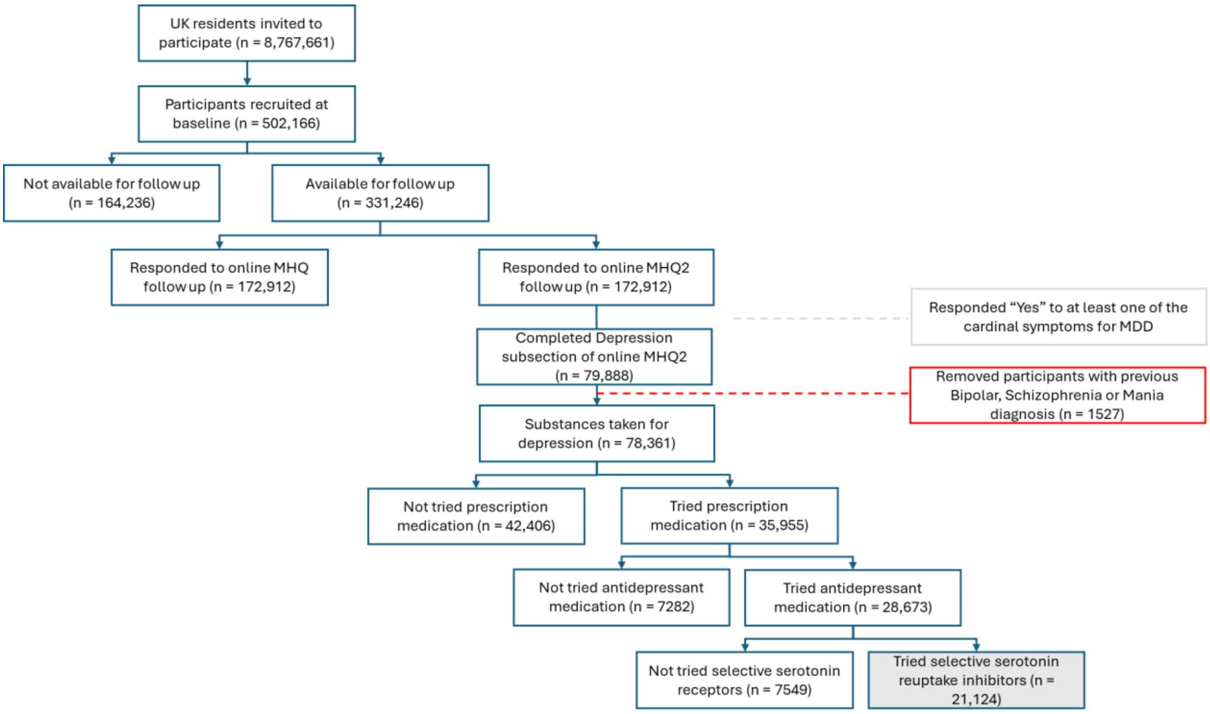

**Figure S1: Flowchart of UK Biobank participants included in antidepressant outcome analyses**

**Decision framework: self-reported SSRI response phenotypes**

This decision framework (detailed in Table S2) accounted for exposure to multiple drugs and aggregated responses across the four SSRIs. Participants' responses were categorised based on their exposure to one, two, three, or four SSRIs:

- Single SSRI Exposure: Participants reporting a positive response ("Y") were classified as responders, while those reporting no response ("N") were classified as non-responders.
- Two SSRI Exposures: Participants with two positive responses ("YY") were classified as responders, and those with two negative responses ("NN") were classified as non-responders. Mixed responses ("YN") were classified as non-responders as most people respond "Y," and "N" are minority, or marked as missing data (NA) if classification was uncertain.
- Three SSRI Exposures: Participants with three positive responses ("YYY") were classified as responders, and those with three negative responses ("NNN") were classified as non-responders. Mixed responses (e.g., "YNN", "YYN") were generally classified as non-responders, particularly if the majority response was negative, or marked as missing data (NA) if uncertain.
- Four SSRI Exposures: Participants with four positive responses ("YYYY") were classified as responders, and those with four negative responses ("NNNN") were classified as non-responders. Mixed responses (e.g., "YNNN", "YYNN", "YYYN") were generally classified as non-responders, particularly if the majority response was negative, or marked as missing data (NA) if uncertain.

**Table S2: Decision framework for defining antidepressant response outcomes among participants using at least one Selective Serotonin Reuptake Inhibitor (SSRI)**

| Number of drugs | Drug specific outcomes | SSRI phenotype | SSRI conservative phenotype |
|---|---|---|---|
| 1 | Y | Y | Y |
| | N | N | N |
| 2 | YY | Y | Y |
| | YN | N | NA |
| | NN | N | N |
| 3 | YYY | Y | Y |
| | YYN | N | NA |
| | YNN | N | N |
| | NNN | N | N |
| 4 | YYYY | Y | Y |
| | YYYN | Y | NA |
| | YYNN | N | N |
| | YNNN | N | N |
| | NNNN | N | N |

### Clinical symptoms and characteristics

**Table S3: Depression clinical characteristics and associated UK Biobank data fields from the MHQ2 questionnaire**

| MDD characteristics | Field ID | Response options[a] | Reference category |
|---|---|---|---|
| Depression possibly related to stressful or traumatic event | 29013 | • No<br>• Yes | Yes |
| Fraction of day affected during worst episode | 29014 | • All day long<br>• Most of the day<br>• About half of the day<br>• Less than half of the day | Most of the day |
| Frequency of depressed days during worst episode | 29015 | • Every day<br>• Almost every day<br>• Less often | Almost every day |
| Brightening of mood in response to positive events during worst episode | 29016 | • No<br>• Yes | Yes |
| Time of day that mood was worse during worst episode. | 29017 | • In the morning<br>• In the evening or at night<br>• Mood did not vary | Mood did not vary |
| Feelings of tiredness during worst episode of depression | 29018 | • No<br>• Yes | Yes |
| Feelings of heaviness in limbs during worst episode of depression | 29019 | • No<br>• Yes | No |
| Change in appetite during worst episode of depression | 29020 | • No change in appetite<br>• Increased appetite<br>• Decreased appetite | Decreased appetite |
| Weight change during worst episode of depression | 29021 | • Gained weight<br>• Lost weight<br>• Both gained and lost some weight during this time<br>• Stayed about the same or was on a diet | Lost weight |
| Difficulty concentrating during worst episode of depression | 29026 | • No<br>• Yes | Yes |
| Feelings of worthlessness during worst period of depression | 29027 | • No<br>• Yes | Yes |
| Feelings of guilt during worst period of depression | 29028 | • No<br>• Yes | Yes |
| Thoughts of death during worst depression episode | 29029 | • No<br>• Yes | Yes |
| Duration of worst depression | 29030 | • Less than a month<br>• Between 1 and 3 months<br>• Over 3 months, but less than 6 months<br>• Over 6 months, but less than 12 months<br>• 1 to 2 years<br>• Over 2 years | Between 1 and 3 months |

| | | | |
|---|---|---|---|
| Impact on normal roles during worst period of depression | 29031 | • A lot<br>• Somewhat<br>• A little<br>• Not at all | A lot |
| Difficulty coping with rejection or negative responses | 29032 | • Yes, caused problems in work/social relationships<br>• Yes, but no problems in work/social relationships<br>• No, this does not sound like me | No, this does not sound like me |
| Lifetime number of depressed periods | 29033 | Continuous number scale binarised to:<br>• Single<br>• Multiple ( >1 episode) | Single |
| Age at first episode of depression | 29034 | Continuous number scale | NA |
| Depression possibly related to childbirth | 29035 | • No<br>• Yes | |
| Age at last episode of depression | 29036 | Continuous number scale | NA |
| Has/did your father ever suffer from? | 20107 | • Hip fracture<br>• Prostate cancer<br>• Severe depression<br>• Parkinson's disease<br>• Alzheimer's disease/dementia<br>• Diabetes<br>• High blood pressure<br>• Chronic bronchitis/emphysema<br>• Breast cancer<br>• Bowel cancer<br>• Lung cancer<br>• Stroke<br>• Heart disease | NA |
| Has/did your mother ever suffer from? | 20110 | | |
| Has/did your father ever suffer from? | 20111 | | |

[a] All data field IDs have “Prefer not to answer” and “I do not know” as a response option

### GWAS summary statistics of Polygenic score analysis

#### Table S4: GWAS summary statistics used to develop five psychiatric disorder-related PGS scores and two antidepressant response-associated trait scores

| Phenotype | Abbr. | PMID | Authors (Ref) | N | Case_N | Control_N |
|---|---|---|---|---|---|---|
| Major Depressive disorder | DEPR | 29700475 (excl. UKB and 23andMe) | Wray et al., 2018 (Wray et al., 2018) | 143,265 | 45,591 | 97,674 |
| Attention Deficit/ Hyperactivity disorder | ADHD | 30478444 | Demontis et al., 2019 (Demontis et al., 2019) | 55,374 | 20,183 | 35,191 |
| Autism | AUTI | 30804558 | Grove et al., 2019 (Grove et al., 2019) | 48,350 | 18,381 | 29,969 |
| Bipolar | BIPO | 31043756 | Stahl et al., 2019 (Stahl et al., 2019) | 147,172 | 9,412 | 137,760 |
| Schizophrenia | SCHI | 29483656 | Pardiñas et al., 2018 (Pardiñas et al., 2018) | 35,802 | 11,260 | 24,542 |
| Antidepressant-non remission | $AD_{non-rem}$ | 35712048 | Pain et al., 2022 (Pain et al., 2022) | 5151 | 3,299 | 1,852 |
| Antidepressant-percentage improvement | $AD_{perc}$ | 35712048 | Pain et al., 2022 (Pain et al., 2022) | 5218 | 5218 | 0 |

### Association of assessment centre

**Table S5: Association  test results between self-reported antidepressant response and UKB Assessment centre**

| Assessment centre | SSRI N = 19516 | | SSRI cons N = 18170 | | Citalopram N = 8335 | | Fluoxetine N = 8476 | | Paroxetine N = 2297 | | Sertraline N = 5883 | |
|---|---|---|---|---|---|---|---|---|---|---|---|---|
| | **OR [95% CI]** | ***p*** | **OR [95% CI]** | ***p*** | **OR [95% CI]** | ***p*** | **OR [95% CI]** | ***p*** | **OR [95% CI]** | ***p*** | **OR [95% CI]** | ***p*** |
| Barts | 1.19 [0.95-1.51] | 0.137 | 1.22[0.93-1.60] | 0.155 | 1.23[0.84-1.80] | 0.282 | 1.02[0.72-1.45] | 0.911 | 0.63[0.32-1.24] | 0.182 | 1.28[0.82-1.98] | 0.281 |
| Birmingham | 1.11[0.92-1.35] | 0.267 | 1.18[0.94-1.47] | 0.149 | 1.06[0.78-1.45] | 0.721 | 1.08[0.80-1.46] | 0.618 | 0.82[0.48-1.41] | 0.471 | 1.05[0.75-1.47] | 0.764 |
| Bristol | 0.98[0.83-1.15] | 0.778 | 1.02[0.84-1.24] | 0.849 | 0.97[0.74-1.27] | 0.802 | 0.99[0.77-1.26] | 0.928 | 1.02[0.63-1.64] | 0.946 | 1.04[0.77-1.40] | 0.815 |
| Bury | 0.94[0.77-1.14] | 0.513 | 0.83[0.65-1.07] | 0.148 | 0.75[0.54-1.05] | 0.098 | 0.99[0.74-1.34] | 0.958 | 0.95[0.53-1.68] | 0.847 | 1.02[0.71-1.46] | 0.912 |
| Cardiff | 1.17[0.95-1.44] | 0.148 | 1.14[0.89-1.46] | 0.312 | 1.06[0.76-1.49] | 0.720 | 1.24[0.91-1.70] | 0.175 | 1.39[0.79-2.44] | 0.257 | 1.29[0.87-1.90] | 0.202 |
| Croydon | 1.08[0.89-1.30] | 0.459 | 1.12[0.90-1.41] | 0.301 | 1.19[0.87-1.63] | 0.264 | 1.05[0.79-1.38] | 0.752 | 0.92[0.53-1.60] | 0.772 | 0.81[0.56-1.18] | 0.277 |
| Edinburgh | 1.01[1.80-1.26] | 0.948 | 0.82[0.61-1.08] | 0.161 | 1.31[0.89-1.92] | 0.171 | 0.79[0.58-1.07] | 0.129 | 0.94[0.51-1.64] | 0.834 | 1.38[0.88-2.15] | 0.158 |
| Glasgow | 1.15[0.93-1.43] | 0.197 | 1.06[0.82-1.38] | 0.660 | 1.17[0.82-1.68] | 0.378 | 1.16[0.86-1.57] | 0.331 | 0.73[0.38-1.40] | 0.348 | 1.00[0.65-1.52] | 0.993 |
| Hounslow | 1.07[1.89-1.30] | 0.464 | 1.08[0.86-1.35] | 0.496 | 0.98[0.71-1.35] | 0.896 | 1.11[0.84-1.46] | 0.467 | 1.14[0.69-1.88] | 0.615 | 1.08[0.76-1.54] | 0.672 |
| Liverpool | 1.04[0.87-1.24] | 0.675 | 1.09[0.89-1.35] | 0.401 | 1.16[0.86-1.56] | 0.322 | 0.92[0.70-1.20] | 0.544 | 0.92[0.56-1.53] | 0.759 | 1.06[0.78-1.46] | 0.696 |
| Manchester | 1.18[0.94-1.48] | 0.144 | 1.08[0.82-1.42] | 0.586 | 1.04[0.71-1.53] | 0.849 | 1.18[0.85-1.63] | 0.328 | 0.71[0.37-1.36] | 0.303 | 1.12[0.75-1.68] | 0.573 |
| Middlesborough | 1.02[0.83-1.25] | 0.853 | 0.97[0.76-1.23] | 0.775 | 0.82[0.57-1.18] | 0.284 | 0.99[0.75-1.32] | 0.967 | 0.86[0.46-1.59] | 0.627 | 1.13[0.80-1.58] | 0.482 |
| Newcastle | 1.07[0.90-1.27] | 0.473 | 1.04[0.84-1.27] | 0.725 | 1.16[0.87-1.56] | 0.315 | 0.90[0.70-1.15] | 0.408 | 1.14[0.69-1.88] | 0.601 | 1.06[0.78-1.45] | 0.710 |
| Nottingham | 0.93[0.77-1.11] | 0.409 | 0.95[0.77-1.18] | 0.653 | 0.93[0.69-1.26] | 0.646 | 0.96[0.74-1.24] | 0.737 | 0.98[0.58-1.67] | 0.946 | 0.92[0.66-1.27] | 0.601 |
| Oxford | 1.13[0.90-1.42] | 0.304 | 1.19[0.71-1.09] | 0.210 | 1.36[0.94-1.95] | 0.099 | 0.88[0.63-1.25] | 0.484 | 0.56[0.26-1.19] | 0.132 | 1.08[0.70-1.67] | 0.720 |
| Reading | 0.96[0.80-1.16] | 0.702 | 0.95[0.824.09] | 0.655 | 1.07[0.79-1.44] | 0.668 | 0.89[0.67-1.16] | 0.383 | 0.67[0.38-1.19] | 0.173 | 1.04[0.74-1.46] | 0.813 |
| Sheffield | 0.97[0.81-1.15] | 0.703 | 0.88[0.64-1.14] | 0.237 | 0.94[0.69-1.29] | 0.703 | 0.74[0.58-0.96] | 0.023 | 1.10[0.67-1.82] | 0.696 | 1.10[0.80-0.50] | 0.568 |
| Stockport | 1.70[0.83-3.47] | 0.146 | 1.83[0.82-4.09] | 0.141 | 1.66[0.59-4.65] | 0.337 | 1.56[0.54-4.56] | 0.413 | 1.54[0.14-17.37] | 0.729 | 1.77[0.45-6.96] | 0.412 |
| Stoke | 0.98[0.78-1.24] | 0.858 | 0.85[0.61-1.14] | 0.280 | 0.89[0.61-1.30] | 0.540 | 0.77[0.54-1.11] | 0.160 | 0.92[0.51-1.68] | 0.789 | 1.02[0.67-1.56] | 0.930 |
| Swansea | 0.93[0.53-1.62] | 0.801 | 0.60[0.27-1.33] | 0.211 | 1.42[0.68-2.95] | 0.346 | 0.57[1.20-1.67] | 0.309 | 1.45E-06[0-4.27E244] | 0.964 | 0.77[0.29-2.03] | 0.592 |
| Wrexham | 0.55[0.19-1.59] | 0.271 | 0.41[0.10-1.73] | 0.224 | 0.36[0.05-2.72] | 0.319 | 1.53[0.47-5.03] | 0.484 | 1.45E-06[0-inf] | 0.988 | 0.32[0.04-2.45] | 0.272 |
| Leeds (ref) | 1.0[1.0-1.0] | NA | 1.0[1.0-1.0] | NA | 1.0[1.0-1.0] | NA | 1.0[1.0-1.0] | NA | 1.0[1.0-1.0] | NA | 1.0[1.0-1.0] | NA |

### Multivariable model Variance Inflation Factors (VIF) analysis

#### Table S6: Variance inflation Factors (VIF) in regression analyses assessing self-reported antidepressant response with sociodemographic factors

| Group | Predictor | Cit | Flu | Par | Ser | SSRI | SSRI_cons |
|---|---|---|---|---|---|---|---|
| All | Sex | 1.03 | 1.03 | 1.05 | 1.03 | 1.03 | 1.03 |
| | Age (scaled) | 1.11 | 1.12 | 1.14 | 1.16 | 1.13 | 1.14 |
| | Ethnic background | 1.03 | 1.03 | 1.03 | 1.04 | 1.03 | 1.03 |
| | Annual income | 1.29 | 1.25 | 1.23 | 1.29 | 1.27 | 1.28 |
| | Highest education | 1.17 | 1.15 | 1.17 | 1.17 | 1.16 | 1.16 |
| | TDI (scaled) | 1.14 | 1.13 | 1.13 | 1.17 | 1.13 | 1.13 |
| | Alcohol status | 1.05 | 1.04 | 1.05 | 1.04 | 1.04 | 1.05 |
| | Smoking status | 1.12 | 1.13 | 1.11 | 1.15 | 1.12 | 1.11 |
| | Self-medicate - drugsandalc | 1.07 | 1.08 | 1.09 | 1.08 | 1.07 | 1.07 |
| Female | Age (scaled) | 1.10 | 1.10 | 1.12 | 1.16 | 1.12 | 1.12 |
| | Annual income | 1.24 | 1.22 | 1.23 | 1.27 | 1.24 | 1.24 |
| | Highest education | 1.15 | 1.14 | 1.16 | 1.16 | 1.14 | 1.15 |
| | TDI (scaled) | 1.12 | 1.10 | 1.12 | 1.15 | 1.11 | 1.11 |
| | Alcohol status | 1.03 | 1.03 | 1.05 | 1.04 | 1.03 | 1.03 |
| | Smoking status | 1.12 | 1.12 | 1.11 | 1.13 | 1.11 | 1.11 |
| | Self-medicate - drugsandalc | 1.06 | 1.06 | 1.08 | 1.07 | 1.06 | 1.06 |
| Male | Age (scaled) | 1.16 | 1.17 | 1.16 | 1.18 | 1.16 | 1.17 |
| | Annual income | 1.39 | 1.32 | 1.30 | 1.34 | 1.32 | 1.32 |
| | Highest education | 1.23 | 1.16 | 1.23 | 1.23 | 1.19 | 1.19 |
| | TDI (scaled) | 1.19 | 1.21 | 1.19 | 1.19 | 1.17 | 1.17 |
| | Alcohol status | 1.05 | 1.05 | 1.06 | 1.03 | 1.03 | 1.04 |
| | Smoking status | 1.14 | 1.14 | 1.16 | 1.16 | 1.13 | 1.13 |
| | Self-medicate - drugsandalc | 1.08 | 1.10 | 1.13 | 1.09 | 1.07 | 1.07 |

*Cit – Citalopram, Flu – Fluoxetine, Par – Paroxetine, Ser – Sertraline, SSRI – Composite-SSRI, SSRI-conservative response phenotype*

**Table S7: Variance inflation Factors (VIF) in regression analyses assessing self-reported antidepressant response with clinical factors**

| Predictor | Cit 1 | Cit 2 | Flu 1 | Flu 2 | Par 1 | Par 2 | Ser 1 | Ser 2 | SSRI 1 | SSRI 2 | SSRI cons | SSRI_cons |
|---|---|---|---|---|---|---|---|---|---|---|---|---|
| Age first episode | 1.36 | 1.35 | 1.33 | 1.33 | 1.49 | 1.48 | 1.36 | 1.36 | 1.34 | 1.35 | 1.38 | 1.38 |
| Age last episode | 1.14 | 1.14 | 1.14 | 1.13 | 1.13 | 1.13 | 1.11 | 1.11 | 1.13 | 1.13 | 1.13 | 1.13 |
| Brightening of mood | 1.38 | 1.33 | 1.38 | 1.33 | 1.34 | 1.28 | 1.37 | 1.31 | 1.36 | 1.31 | 1.37 | 1.31 |
| Change in appetite | 3.16 | - | 3.46 | - | 4.08 | - | 2.92 | | 3.14 | | 3.06 | - |
| Difficulty coping rejection | 1.29 | 1.27 | 1.28 | 1.28 | 1.29 | 1.28 | 1.30 | 1.28 | 1.27 | 1.26 | 1.28 | 1.27 |
| Feelings of guilt | 1.39 | 1.39 | 1.38 | 1.39 | 1.43 | 1.41 | 1.44 | 1.43 | 1.40 | 1.40 | 1.42 | 1.42 |
| Feeling of heavy limbs | 1.12 | 1.11 | 1.12 | 1.11 | 1.12 | 1.12 | 1.11 | 1.11 | 1.11 | 1.10 | 1.12 | 1.11 |
| Feeling worthlessness | 1.47 | 1.46 | 1.44 | 1.44 | 1.52 | 1.54 | 1.39 | 1.40 | 1.45 | 1.45 | 1.47 | 1.48 |
| Roles impacted | 1.40 | 1.35 | 1.34 | 1.31 | 1.48 | 1.41 | 1.39 | 1.34 | 1.35 | 1.31 | 1.37 | 1.33 |
| Duration of worst episode | 1.22 | 1.19 | 1.24 | 1.22 | 1.33 | 1.28 | 1.22 | 1.19 | 1.19 | 1.17 | 1.20 | 1.18 |
| Fraction of day affected | 1.93 | 1.46 | 1.80 | 1.43 | 1.81 | 1.45 | 2.10 | 1.45 | 1.91 | 1.43 | 1.92 | 1.44 |
| Frequency of depressed days | 1.86 | - | 1.68 | - | 1.76 | - | 1.97 | - | 1.82 | - | 1.83 | - |
| Thoughts of death | 1.14 | 1.15 | 1.15 | 1.15 | 1.21 | 1.20 | 1.15 | 1.14 | 1.14 | 1.14 | 1.15 | 1.15 |
| Weight change | 3.22 | 1.20 | 3.51 | 1.22 | 4.06 | 1.27 | 2.95 | 1.23 | 3.17 | 1.19 | 3.10 | 1.20 |
| Episodes | 1.35 | 1.35 | 1.36 | 1.36 | 1.43 | 1.40 | 1.31 | 1.31 | 1.35 | 1.35 | 1.40 | 1.40 |
| Depression related to trauma | 1.11 | 1.10 | 1.10 | 1.10 | 1.16 | 1.15 | 1.10 | 1.09 | 1.11 | 1.10 | 1.11 | 1.11 |
| Family history | 1.03 | 1.03 | 1.03 | 1.02 | 1.06 | 1.05 | 1.03 | 1.03 | 1.02 | 1.02 | 1.02 | 1.02 |
| Age | 1.15 | 1.14 | 1.13 | 1.12 | 1.21 | 1.19 | 1.17 | 1.16 | 1.13 | 1.12 | 1.13 | 1.12 |
| Sex | 1.08 | 1.07 | 1.10 | 1.09 | 1.15 | 1.12 | 1.10 | 1.08 | 1.09 | 1.07 | 1.09 | 1.08 |

*Cit – Citalopram, Flu – Fluoxetine, Par – Paroxetine, Ser – Sertraline, SSRI – Composite-SSRI, SSRI-conservative response phenotype*

### Antidepressant response and exposure

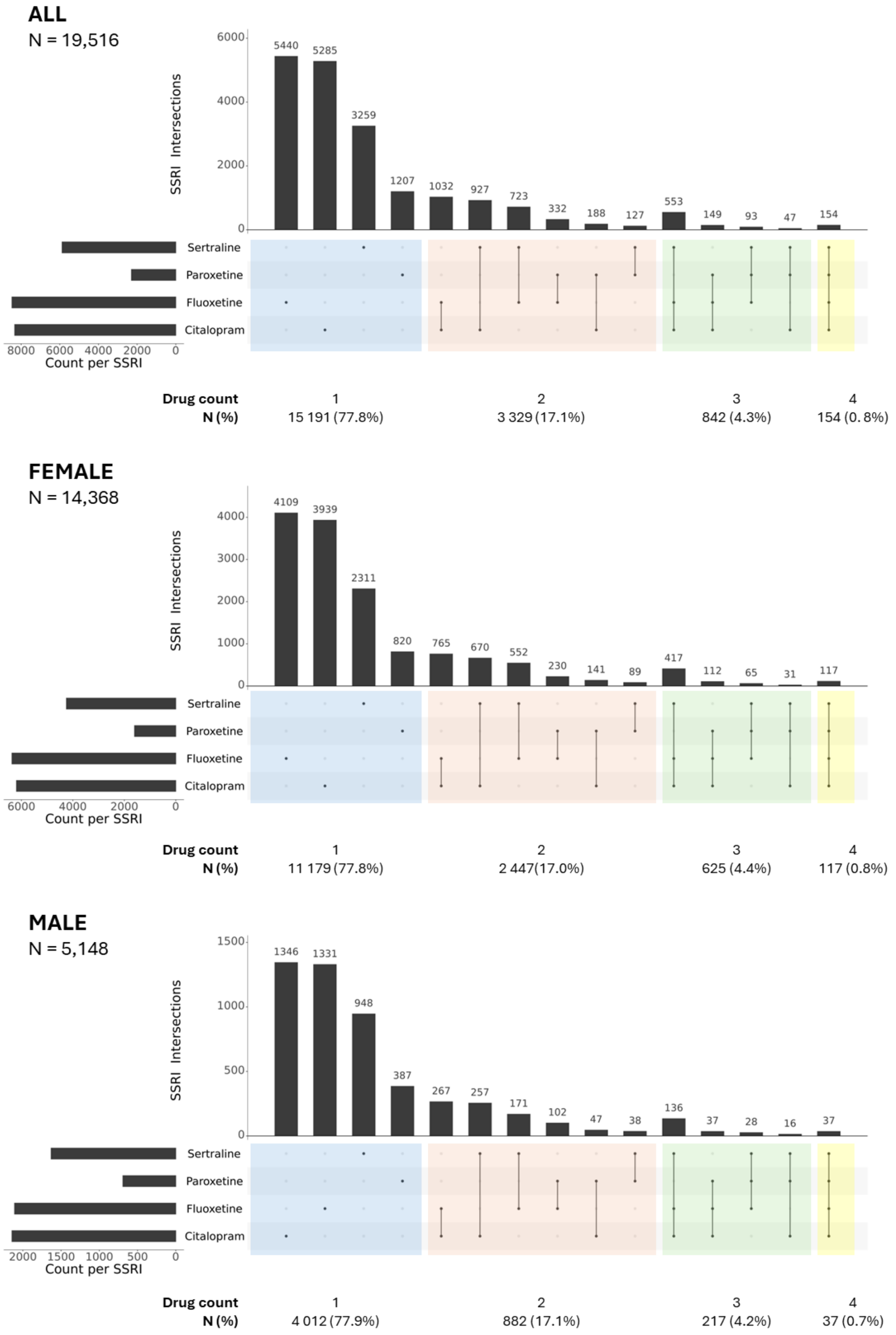

**Figure S2: Upset plot showing number of participants trying different SSRI combinations in the UK Biobank.**

Counts derived from those who responded "Yes", "No", to whether the drug (Citalopram, Fluoxetine, Paroxetine and Sertraline) had made them feel better. *SSRI – Selective serotonin reuptake inhibitors.*

**Table S8: Self-reported SSRI response rate comparison across drugs**

| Group | Drug | Yes (%) | No (%) |
|---|---|---|---|
| All | SSRI | 79. 6 | 20.4 |
| Females | SSRI | 80.7 | 19.3 |
| Males | SSRI | 76.5 | 20.4 |

**Table S9: Drug-specific response proportions and chi-squared test results**

| Group | Drug | Yes (%) | No (%) | χ2 stat | Df | Std res: No | Std res: Yes | *P* |
|---|---|---|---|---|---|---|---|---|
| All | Citalopram | 82.8 | 17.2 | 73.28 | 3 | -7.89 | 7.89 | 8.483e-16 |
| | Fluoxetine | 78.2 | 21.8 | | | 5.08 | -5.08 | |
| | Paroxetine | 76.8 | 23.2 | | | 4.01 | -4.01 | |
| | Sertraline | 79.8 | 20.2 | | | 0.38 | -0.38 | |
| Females | Citalopram | 83.9 | 16.1 | 57.16 | 3 | -7.05 | 7.05 | 2.37e-12 |
| | Fluoxetine | 80.2 | 19.8 | | | 2.09 | -2.09 | |
| | Paroxetine | 77.3 | 22.7 | | | 4.08 | -4.08 | |
| | Sertraline | 79.6 | 20.4 | | | 2.82 | -2.82 | |
| Males | Citalopram | 79.4 | 20.6 | 48.95 | 3 | -3.51 | 3.51 | 1.34e-11 |
| | Fluoxetine | 71.8 | 28.2 | | | 6.54 | -6.54 | |
| | Paroxetine | 75.6 | 24.4 | | | 0.80 | -0.80 | |
| | Sertraline | 80.3 | 19.7 | | | -3.82 | 3.82 | |

**Table S10: Sample distribution of inferred metabolizer status across selective serotonin reuptake inhibitors (SSRI).**

| Metaboliser status | SSRI N (%) | SSRI-cons[a] N (%) | Citalopram[b] N (%) | Fluoxetine[b] N (%) | Paroxetine[b] N (%) | Sertraline[b] N (%) |
|---|---|---|---|---|---|---|
| Poor | 443 (2.3) | 411 (2.3) | 197 (2.4) | 197 (2.4) | 47 (2.1) | 128 (2.2) |
| Normal | 7 507 (39.5) | 6 984 (39.5) | 3 225 (39.7) | 3 258 (39.5) | 853 (382) | 2 270 (39.7) |
| Intermediate | 4 943 (26.0) | 4 596 (26.0) | 2 143 (26.4) | 2 144 (26.0) | 584 (26.1) | 1 480 (25.9) |
| Rapid | 5 155 (27.1) | 4 806 (27.2) | 2 162 (26.6) | 2 247 (27.3) | 637 (28.5) | 1 536 (26.9) |
| Ultra rapid | 930 (4.9) | 862 (4.9) | 391 (4.8) | 393 (4.8) | 111 (5.0) | 293 (5.1) |
| Indeterminate | 14 (0.1) | 13 (0.1) | 9 (0.1) | 5 (0.1) | 2 (0.1) | 4 (0.1) |
| Total N | 18 992 | 17 672 | 8 127 | 8 224 | 2 234 | 5 711 |

[a] SSRI-cons – SSRI-conservative response phenotype. [b] Sample sizes across SSRIs do not add to the total SSRI sample size as some participants reported taking more than one antidepressant.

### Phenotypic variance explained of self-reported antidepressant non-response by psychiatric and antidepressant response PGS in UK Biobank

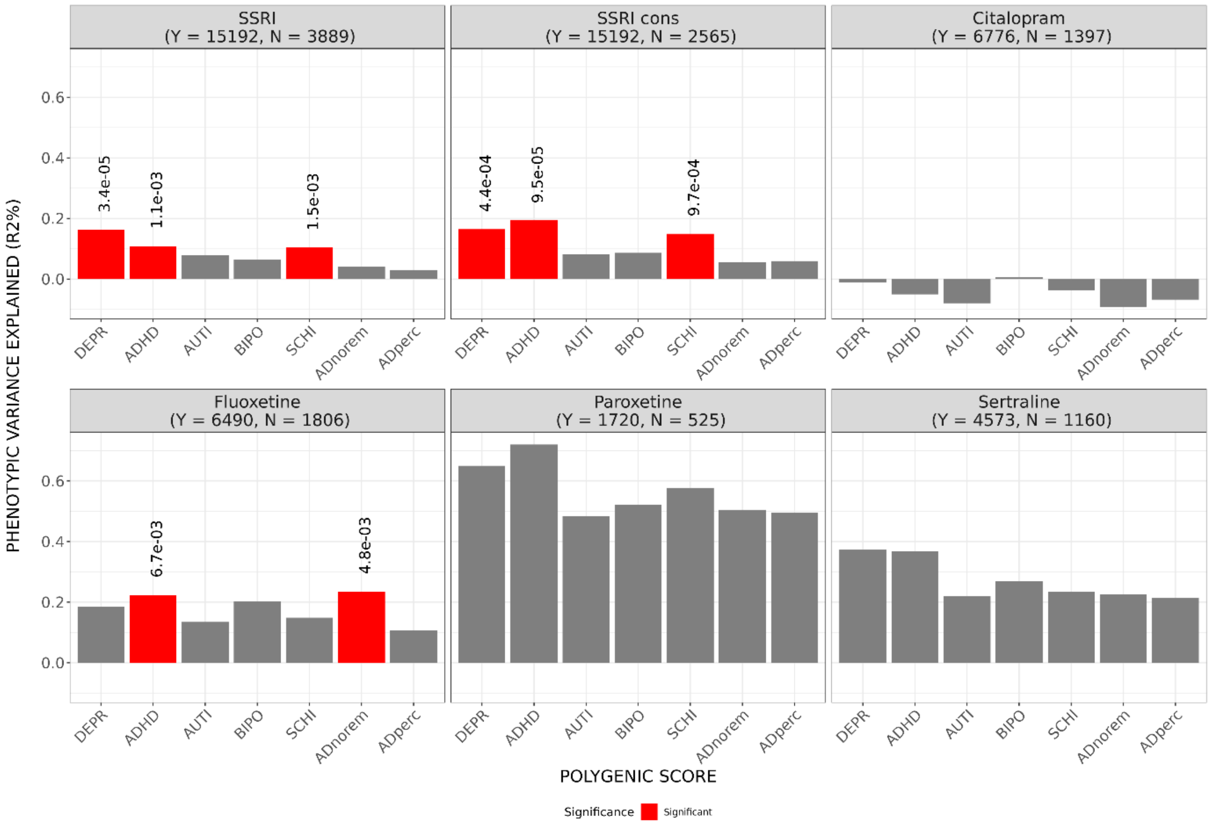

**Figure S3: Phenotypic variance explained (R2%) of self-reported antidepressant non-response by psychiatric and antidepressant response PGS in UK Biobank.** Variance of self-reported antidepressant non-response explained by various mental health condition and treatment polygenic scores between (PGS) for different antidepressants: SSRIs and specific SSRIs (Citalopram, Fluoxetine, Paroxetine, Sertraline). PGS include DEPR: Depression, ADHD: Attention Deficit Hyperactivity Disorder, AUTI: Autism, BIPO: Bipolar Disorder, SCHI: Schizophrenia. ADperc: Percentage improvement, ADNorem: AD non-remission. Multiple testing correction across PGS, within each SSRI P < 0.007.
