## Supplementary materials 2 for "Sociodemographic, clinical, and genetic factors associated with self-reported antidepressant response outcomes in the UK Biobank"

Table S1: Univariable analysis assessing sociodemographic variables associated with self-reported a
Table S2: Multivariable analysis assessing sociodemographic variables associated with self-reported
Table S3: Univariable analysis assessing clinical variables associated with self-reported antidepressa
Table S4: Multivariable analysis assessing clinical variables associated with self-reported antidepress
Table S5: CYP2C19 metaboliser status associated with self-reported antidepressant response in a su
Table S6: Polygenic risk scores associated with self-reported antidepressant response in a subsampl

ntidepressant response in a subsample of the UK Biobank. Odds ratios, 95% confidence intervals and p-values est
antidepressant response in a subsample of the UK Biobank. Odds ratios, 95% confidence intervals and p-values e
nt response in a subsample of the UK Biobank. Odds ratios, 95% confidence intervals and p-values estimated fror
ant response in a subsample of the UK Biobank. Odds ratios, 95% confidence intervals and p-values estimated frc
osample of the UK Biobank. Odds ratios, 95% confidence intervals and p-values estimated from regression analys
e of the UK Biobank. Odds ratios, 95% confidence intervals and p-values estimated from regression analyses of an

timated from regression analyses of antidepressant non-response outcomes.
stimated from regression analyses of antidepressant non-response outcomes.
